# Maternal Mortality and Women’s Political Participation

**DOI:** 10.1101/19000570

**Authors:** Sonia Bhalotra, Damian Clarke, Joseph Gomes, Atheendar Venkataramani

## Abstract

We show that large declines in maternal mortality can be achieved by raising women’s political participation. We estimate that the recent wave of quotas for women in parliament in low income countries has resulted in a 9 to 12% decline in maternal mortality. Among mechanisms are that gender quotas lead to an 8 to 10% increase in skilled birth attendance, a 6 to 12% increase in prenatal care utilization and a 4 to 11% decrease in birth rates.

**JEL codes:** I14, I15, O15.

## 1 Introduction

Maternal mortality, defined as the death of women within 42 days of childbirth, remains a looming global health problem well into the 21^st^ century. It is estimated to account for 830 deaths per day, and more than 216 deaths per 100,000 live births globally (Ceschia and Horton, 2016). In sub-Saharan Africa, the maternal mortality ratio (MMR) exceeds the rate in developed countries a century ago (Alkema et al., 2016; Loudon, 1992).^1^ Moreover, maternal mortality is only the tip of an iceberg, the mass of which is maternal morbidity.

Persistence of high rates of maternal mortality is striking given that the knowledge and technology needed to dramatically reduce it have been available for nearly a century, and the costs of intervention are relatively small (Cutler, Deaton and Lleras-Muney, 2006; Loudon, 1992). There remains far from universal coverage of reproductive health services in low income countries. Since 99% of maternal mortality occurs in developing countries, a natural explanation may be that low income has constrained progress. However, while income displays a positive association with each of female and male life expectancy, it exhibits only a weak relationship with the ratio of female to male life expectancy, a proxy for excess deaths of women associated with reproduction (Appendix Figure B1).^2^ This suggests other factors at play.^3^ We investigate the hypothesis that, among other factors, are gendered policy preferences. In particular, that addressing maternal mortality has been a low priority in male-dominated parliaments.

Maternal mortality has declined rapidly in the last two decades, but there was massive variation in rates of decline.^4^ We leverage this variation to investigate the hypothesis that political will plays a significant role, and that women have greater political will (and/or efficacy) than men for maternal mortality reduction. The broad stylized facts line up with our hypothesis: since 1990 MMR has shown an unprecedented fall of 44%, a period in which the share of women in parliament has risen unusually rapidly, from under 10% to more than 20% (Figure 1a). We study whether these trends are causally related.

**Figure 1:**
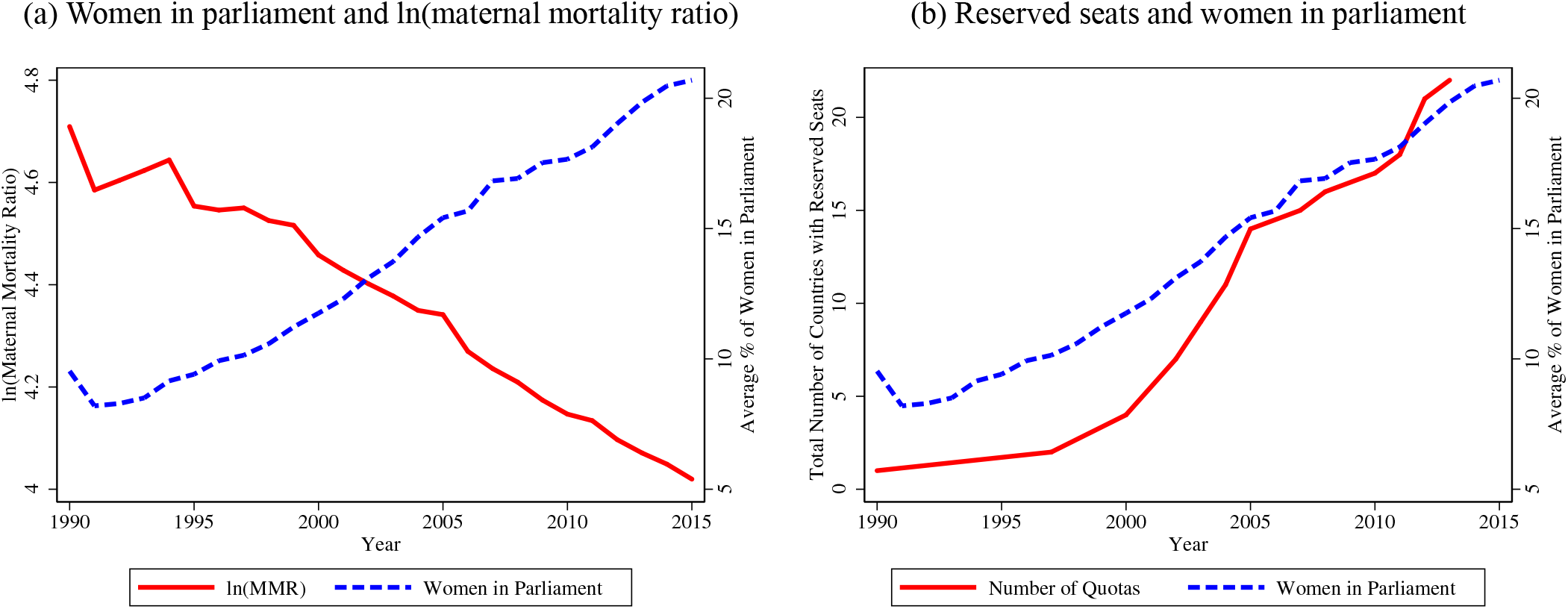
Trends in gender quotas, women in parliament and maternal mortality. Notes: Raw trends in number of countries with parliamentary gender quotas, the percentage of women in parliamentary seats and the log of the maternal mortality ratio. Data sources are provided in the Data Appendix. The sample is a global sample of 174 countries for which we have annual data through 1990–2015.

The share of women in parliament has been increasing at an increasing rate but smoothly, making it hard to isolate its effects from those of other gradually evolving trends. We address this problem by exploiting the abrupt legislation of parliamentary gender quotas sweeping through developing countries since the early to mid 1990s. The identifying assumption is that, conditional on country and year fixed effects, the timing of quota implementation is quasi-random. Figure 1b shows that trends in women’s share in parliament track trends in quota coverage. We combine country-specific dates of quota implementation with the first annualized estimates of MMR across countries, released in 2015 to generate a global country-year panel for 1990–2015. Our strategy is to estimate event study style regressions that show trends in MMR pre- and post-quota adoption (Jacobson, LaLonde and Sullivan, 1993; Goodman-Bacon, 2018).

Our estimates show that passage of parliamentary gender quotas leads to an immediate 5 to 6 percentage point (55 to 66%) increase in the share of parliamentary seats held by women, and a 9 to 12% decrease in the maternal mortality ratio. The effects of quotas are increasing in time since implementation, the share of seats reserved, and in pre-intervention maternal mortality rates. There is no evidence of differential pre-trends in the outcomes. Moreover, the estimates are robust to using an alternate data source on MMR, changes in sample composition, changes in covariates (including adjustment for predictors of quota implementation identified in the political science literature), and addressing the degree of potential bias from any additional unobserved confounding in an IV setup using the method of Conley, Hansen and Rossi (2012). We also address measurement issues, showing results for a balanced sample, using alternative measures of MMR, and using alternative inference procedures.

Investigating mechanisms that may link an increasing share of women in parliament to lower maternal mortality, we find evidence in favor of investments in key low-cost medical inputs. Gender quotas result in a 6.9 to 8.2 percentage point (8.3–9.8%) increase in skilled birth attendance and a 5.1 to 9.8 percentage point (6.2–11.8%) increase in antenatal care utilization. The WHO recommends universal access to these inputs, and they are widely promoted as tools for maternal mortality reduction (WHO, 2014; Jamison et al., 2013).^5^ We also find a 4–10% decline in birth rates and some tendency for an increase in girls’ education following gender quotas, both of which previous research has shown to be associated with lower maternal mortality. We also tested whether quotas lead to increases in the share of development assistance for maternal health from international donors, as this is thought to have played a critical role in the world’s response to the Millennium Development Goals (one of which was MMR reduction) since 2000 (Dieleman et al., 2016), but we find no evidence of this. We find no discernible impact of gender quotas on GDP, or on health expenditure as a share of GDP. Importantly, the relationship between MMR and gender quotas is not sensitive to conditioning upon these variables.

It is notable that income has no impact on the share of women in parliament, but it has a significant impact on MMR. A 1% increase in GDP results in a 0.5% reduction in MMR. A crude back-of-the-envelope calculation assuming log-linearity, and holding democracy and quotas constant suggests that to achieve the estimated 9–12% reduction in MMR due to quota adoption (on the same sample), GDP would have to increase by about 18–24%. Income is potentially endogenous so this is only a crude calculation but note that we test whether GDP changes in response to quotas and we find it does not.

These results suggest that the operative mechanism linking parliamentary gender quotas to MMR reduction may have been a more efficient allocation of existing resources to reflect refreshed priorities, and improved allocative efficiency. We probe this by examining whether gender quotas were associated with detrimental effects on other health outcomes. We find no significant impact of quotas on adult male mortality, a crude analogue of maternal mortality. We also find no impact on tuberculosis mortality, a highly prevalent disease in low income countries that mostly affects adults in their (re)productive years and that has been equally burdensome for men and women. We find some evidence that gender quotas result in a fall in infant mortality for girls, but not boys. Overall, the evidence indicates that women parliamentarians are more effective in targeting women’s health than at targeting health in general and, further, that they appear to improve allocative efficiency.

To summarize, using longitudinal cross-country data across a period of 25 years that encompasses periods of dramatic decline in maternal mortality, we provide compelling new evidence that raising women’s political participation can have substantial impacts on maternal mortality. Overall, quotas led to a 13% decline in maternal mortality within 10 years of implementation, comparing favorably to the 44% decline in MMR globally that occurred over a 25 year period.^6^ The importance of women’s political participation is underscored by a dose-response relationship: countries that reserved 20-30% of parliamentary seats, close to the internationally recommended target, experienced an 18% decline in MMR. We also find that the impact of gender quotas on MMR is increasing in the baseline rate of MMR, consistent with the “low-hanging fruit” argument, or diminishing returns.

Efforts to reduce maternal mortality over our study period have focused on raising access to trained birth assistance, prenatal and antenatal care, contraception and women’s education (Grépin and Klugman, 2013; Kruk et al., 2016). There has been no recognition among policy makers of the potential relevance of the political economy of resource allocation influencing these inputs. However, the 6 to 9 percentage point increase in birth attendance and the 5 to 9 percentage point increase in prenatal care that we demonstrate flow from quota passage compare well with the 12 and 13 percentage point increases achieved through the recent 25 years.

Our study makes two key contributions. We are the first to propose that gender quotas can be an effective policy tool for maternal mortality reduction. This is important because (i) the broader evidence on the success of quotas is mixed (Coate and Loury, 1993; Besley et al., 2017; Pande and Ford, 2012; Niederle, 2016), and (ii) MMR has been difficult to bring down. Regarding the latter, the decline in MMR of 44% since 1990 fell well short of the Millennium Development Goal (MDG) target decline of 75% (Hogan et al., 2010; Kassebaum et al., 2014), and yet the new Sustainable Development Goals (SDGs) have set a higher target. This is a clear flag that some policy innovation is needed. Our results suggest that women leaders are more effective than men in implementing the known recipes for success in this domain.

Second, we provide possibly the first systematic analysis of the impacts of the recent wave of implementation of gender quotas across countries.^7^ Our findings cohere with previous evidence that increasing the share of women politicians influences policy choices in favor of public goods or policies that align with the preferences of women (Chattopadhyay and Duflo, 2004; Taylor-Robinson and Heath, 2003; Swers, 2005; Clots-Figueras, 2012; Kose, Kuka and Shenhav, 2016). Experimental research showing that women have different preferences from men provides a behavioural underpinning to these results (Niederle, 2016).

Reducing maternal mortality is of both intrinsic and functional value, as it favorably influences women’s human capital attainment, employment, and growth (Albanesi and Olivetti, 2016, 2014; Jayachandran and Lleras-Muney, 2009; Bloom, Kuhn and Prettner, 2015).^8^ A broad stream of research has documented the importance of population health for economic growth, via life expectancy and human capital accumulation (Soares, 2005; Weil, 2007; Ashraf, Lester and Weil, 2009; Shastry and Weil, 2003; Bloom, Canning and Sevilla, 2004; Lorentzen, McMillan and Wacziarg, 2008; Aghion, Howitt and Murtin, 2010).

The rest of this paper is organized as follows. In section 2 we provide a discussion of the implementation of parliamentary gender quotas. In section 3 we describe the data used in this paper, and in section 4 we lay out the empirical strategy. We present results in section 5, and discuss the mechanisms underlying these results in section 6. We briefly conclude in section 7.

## 2 Gender Quotas – Background

In response to an active civil society movement and rising awareness of women’s rights, in 1990 the UN Economic and Social Council set a target of 30% female representation in decision making bodies by 1995 (Pande and Ford, 2012; UN Women, 1995). The passage of gender quotas followed this and accelerated after the unanimous signing of the Beijing Platform for Action by all UN delegates at the Fourth World Conference on Women in 1995 (Inter-Parliamentary Union, 2015; Chen, 2010; Krook, 2010). Since 1990, 22 countries in sub-Saharan Africa, the Middle East, and South and East Asia have implemented constitutionally protected quotas reserving seats in parliament for women. The geographic spread and trend in gender quotas is described in Figures A1 and A2.^9^ We observe an uptick in quotas particularly after year 2000, with these being most prevalent in Sub-Saharan Africa. The distribution of the share of seats reserved for women by legislative changes is described in Figure A3, the median (mean) gender quota is 21% (20%). In section 5.2, we identify country-specific predictors of quota implementation.

While the main focus of our study is reserved seat quotas, since 1990 the number of countries with candidate list quotas for women has also risen sharply (from 1 to 46). As candidate quotas have a different geographical spread, tending to have been passed in middle and higher income countries rather than in low income countries (see Figure A1), we analyse them separately.

Casual inspection suggests support for our hypothesis that reserved seat quotas are associated with MMR decline. Comparing country pairs with similar GDP per capita in 1990, selecting one which implemented reserved seat quotas before 2010 and one which did not, we found that the quota-implementing country typically witnessed a larger decline in maternal mortality in 1990–2010. Thus, Burundi did better than Malawi, Kenya did better than Zimbabwe and Niger did better than the DRC.

## 3 Data

We merge quota dates with MMR data and the estimation sample contains (at most) 174 countries, through 1990–2015. Summary statistics are in Table A1. The data on country-specific adoption of parliamentary gender quotas up until 2005 are from Dahlerup (2005). We updated these using the Global Database of Quotas for Women. The data include information on the date of passage and the share of seats reserved for women. We merge these data to a comprehensive country-year panel on the share of women in parliament aggregating information from multiple sources, namely the World Development Indicators (WDI), the UN Millennium Development Goals (MDG) Indicators and the ICPSR dataset compiled by Paxton, Green and Hughes (2008).

For maternal mortality, we use estimates recently made available from the United Nations Mortality Estimation Inter-Agency Group (MMEIG) containing data for 1990–2015 for as many as 183 countries. These are widely considered the best MMR estimates to date, as they address known measurement difficulties in survey and vital statistics data on maternal mortality using Bayesian methods applied to multiple, complementary data sources including vital statistics, special inquiries, surveillance sites, population-based household surveys and census files (Alkema et al., 2016, 2017). The world distribution of average MMR for the period of 1990–2015 from these data is represented in Appendix Figure A4. Prior to the release in 2015 of these MMR data, there were no annual time series for a comprehensive set of countries. This has no doubt contributed to maternal mortality being vastly understudied relative to, say, infant mortality. As these data include measures of MMR that are estimated, we conduct a sensitivity check that allows for this in inference. We also show that our results hold using an MMR measure from an alternate dataset, the Demographic and Health Surveys (DHS), in which consistent estimates over time are available for a restricted sample of countries. The data and checks are discussed further in section 5.2.

We also use data on a range of time-varying controls, intermediate outcomes and placebo outcomes. Most of these data are compiled from large cross-country databases including the World Bank’s World Development Indicators (WDI) database. For some of these variables we have observations for only a subset of years. Appendix A provides details of all the variables used, and Appendix Table A1 provides the summary statistics.

## 4 Empirical Strategy

We exploit the staggered timing of the implementation of quotas across countries, looking to identify causal effects from significant breaks in the outcome series following implementation. Research designs working in such a setting often employ the single coefficient difference-in-differences (DD) estimator. As shown in Goodman-Bacon (2018), this is only strictly valid when treatment occurs once, between the pre- and the post-period, generating fixed treated and control units. When treatment varies over time, arriving in some regions after others, there are in fact multiple experiments. Already treated units can act as controls for later treated units because their treatment status does not change. However, if there are changes in treatment effects over time, these get subtracted from the DD estimate, biasing the single coefficient estimator away from the true treatment effect. This is not a problem with the underlying design but, rather, with the restriction to a single coefficient. For this reason, our preferred estimates for our main outcomes come from flexible event study models (Jacobson, LaLonde and Sullivan, 1993).

Our main event study model is as follows:

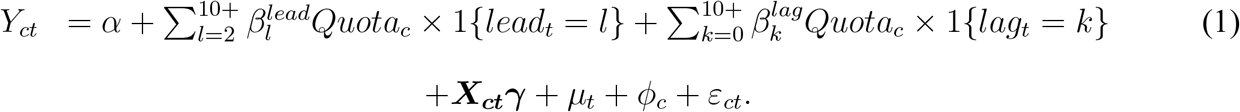

The variation is across country *c* and year *t*. The outcome *Y*_*ct*_ is, first, the proportion of women in parliament, allowing us to estimate how effective gender quotas are at increasing representation. What we may think of as a second-stage outcome but in fact is estimated as a reduced form outcome is the natural logarithm of the maternal mortality ratio. *Quota*_*c*_ is 1 if a country ever adopted a quota, and this is interacted with a full set of leads and lags with respect to the year the quota was adopted. We include 10 lags and leads, the tenth term including all years greater than 10, and the first lead is omitted as the base category. We include country and year fixed effects (*ϕ*_*c*_ and *µ*_*t*_ respectively), and cluster standard errors at country level (Bertrand, Duflo and Mullainathan, 2004).

The *β*^*lag*^ coefficients capture the impacts of interest and the *β*^*lead*^ coefficients partially test the identifying assumption of no differential pre-trends. The *β*^*lead*^ coefficients provide only a partial test of the identifying assumptions, as to estimate unbiased parameters we require parallel trends between treated and non-treated units *in the absence of treatment*. Parallel pre-trends provide support for this assumption, but cannot be used to test what would have happened at the time of the reform had the reform not been implemented (Kahn-Lang and Lang, 2018).

As income and democracy are potentially correlated with both quotas and MMR, we include log GDP per capita and democracy score as time-varying covariates ***X***_***ct***_. However as these controls are potentially endogenous, we show results with and without them. We implement a number of other checks, including testing robustness to including potential predictors of quota legislation as controls in both estimating equations, and estimating bounds on IV estimates that allow for failure of the exclusion restriction (Conley, Hansen and Rossi, 2012).

We also estimate a parametric difference in difference (DD) specification where the independent variable is defined as one for all years following the implementation of a quota for implementing countries, and zero before. It is set to zero for all countries that do not implement quotas in the sample period. In a specification check, we drop the 47 high income countries from the sample so that the control group is more homogeneous. In general, though, we retain in the sample the many (high and low income) countries that do not pass quotas in the sample period, as this expands the set of good comparisons available to identify trends (Borusyak and Jaravel, 2017). Borusyak and Jaravel (2017) show that the DD specification will tend to estimate an average of treatment effects that over-weights short-run effects and under-weights long-run effects. Since the event study plots suggest that treatment effects increase with time since the quota event, in our setting the DD model will produce conservative estimates.

Since the best available country-year panel data on MMR are estimated (due to gaps in vital statistics) and are published with uncertainty bounds (Alkema et al., 2016), we show estimates using a double-bootstrap procedure re-sampling over the uncertainty intervals to calculate the standard errors. About 76% of the country-year observations in the MMR data that we use in the main estimates are original survey data points, the remaining 24% being imputed. An important further check we conduct to address this potential concern is that we re-estimate the model using a different data source—the DHS—for which systematic and comparable survey data on MMR are available, albeit for a smaller sample of countries.

As the countries in the sample vary considerably in population size, we re-estimated the equation weighting by this. Solon, Haider and Wooldridge (2015) argue that this affords a test of model mis-specification. Since MMR varies considerably across countries, proportional changes implied by using logarithms will exaggerate achievements in countries with lower baseline rates (Deaton, 2006). We therefore replaced the logarithm with the level of MMR as a robustness exercise. We also investigate intensive margin effects, exploiting variation in quota size, and we investigate effects by duration and by baseline rates of MMR.

To understand the scope of women’s influence on health outcomes, we replace MMR with mortality for adult males (age 15–60), roughly mirroring the age profile of MMR.^10^ We also produce estimates for tuberculosis mortality and infant mortality as these affect both genders.^11^ Since the TB data had the occasional zero, we use the inverse hyperbolic sine transformation rather than the log transformation for this outcome. For completeness, we show results for adult female mortality as well. After presenting the main results we investigate the impact of quotas on a series of intermediate outcomes with a view to identifying mechanisms.

## 5 Results

### 5.1 Principal Estimates

Estimates of Equation 1 are in Figure 2. Panel A shows a discrete jump in women’s parliamentary representation in the year after quotas are implemented. Panel B shows a break in the coefficient series, with maternal mortality falling more rapidly in quota implementing countries. The drop is visually apparent in the year after implementation, and becomes statistically significant two years after quotas are introduced and is then sustained at an increasing rate. The lead coefficients allay concerns about endogeneity of policy adoption, as they are quite tightly centred around zero.

**Figure 2:**
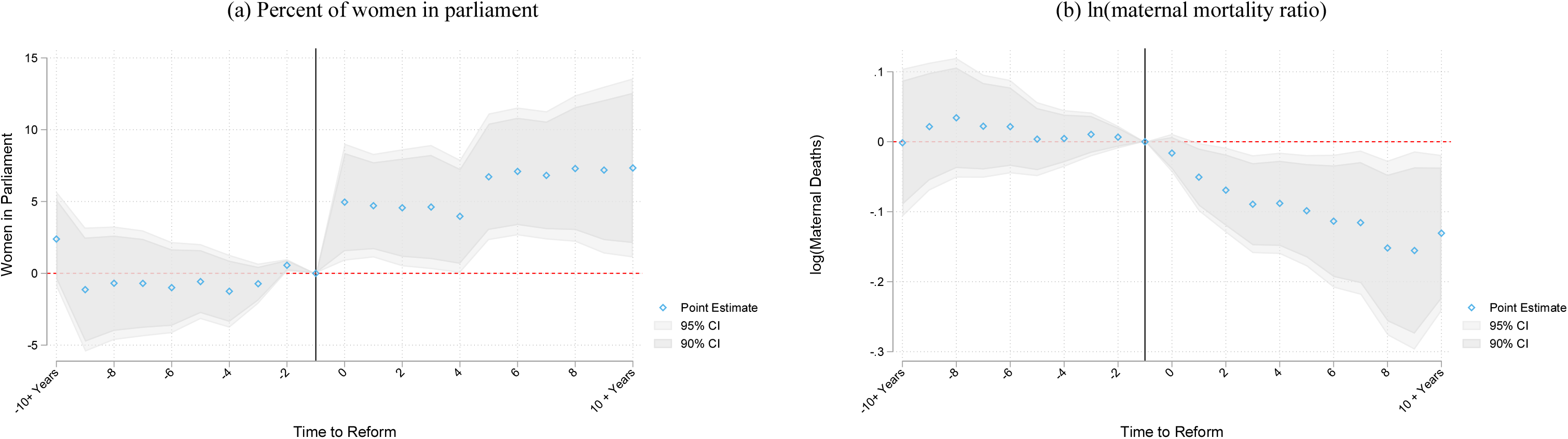
Gender quotas: Event studies for women in parliament and maternal mortality. Notes: Point estimates of the lag and lead terms in the event study specification described in equation 1 are presented, along with their 90% and 95% confidence intervals. Estimates are conditional on country and year fixed effects, the natural logarithm of per capita GDP, and indicators for levels of the Polity IV democracy index. Time periods greater than 10 years from the reform date are displayed as a single “10 +” indicator. Standard errors are clustered by country. The omitted base category is taken as 1 year prior to the reform, indicated by the solid vertical line.

The single coefficient difference-in-differences models are useful complements to the plots as they allow us to report an effect size, which also enables comparison with effect sizes in other studies that rely upon the DD specification. See Table A2. Following Figure 2, we allow a one year lag for the share of women in parliament, and an additional year for impacts on maternal mortality. As the share of women can only change at the next election, we identified for every country, the years between quota legislation and election. The mode and median are zero years, the mean is 1.3. Once women are in parliament, it is plausible that it takes (at least) one year for any changes they induce to have discernible population-level impacts on maternal mortality.^12^ The point estimates indicate statistically significant effects of gender quotas on the proportion of women in parliament of 5–6 percentage points which, relative to the average in 1985–1990 of 9%, represents a 55 to 66% increase.^13^ We also identify a substantial reduction in MMR of 9–12%. In considering the effect size, we avoid linear extrapolation as the effects are non-linear in quota size. This is discussed below.

### 5.2 Robustness Checks and Extensions

#### Sample, functional form, weights, endogenous surveillance, clustering

Dropping high income countries and re-estimating produces essentially identical estimates (Figure 3a). To assess sensitivity of our estimates to compositional change, we dropped the 7 countries that passed quotas after 2005 to create a balanced sample. The estimates again are essentially unchanged (Figure 3b). Figure 3c and Table A2 also shows that the estimates are robust to using level rather than log MMR. Results with population weights are provided in Figure 3d. Since China and India are outliers in population size, the weighted estimates exclude them (China implemented quotas, India did not). So as to isolate changes ensuing from weighting from changes associated with removing these countries, we also show unweighted estimates on the reduced sample (Figure 3e). The point estimates in a single term DD model are larger with China and India excluded, and again larger when weighted. However, all changes in the sequence are not statistically meaningful (Table A2).

**Figure 3:**
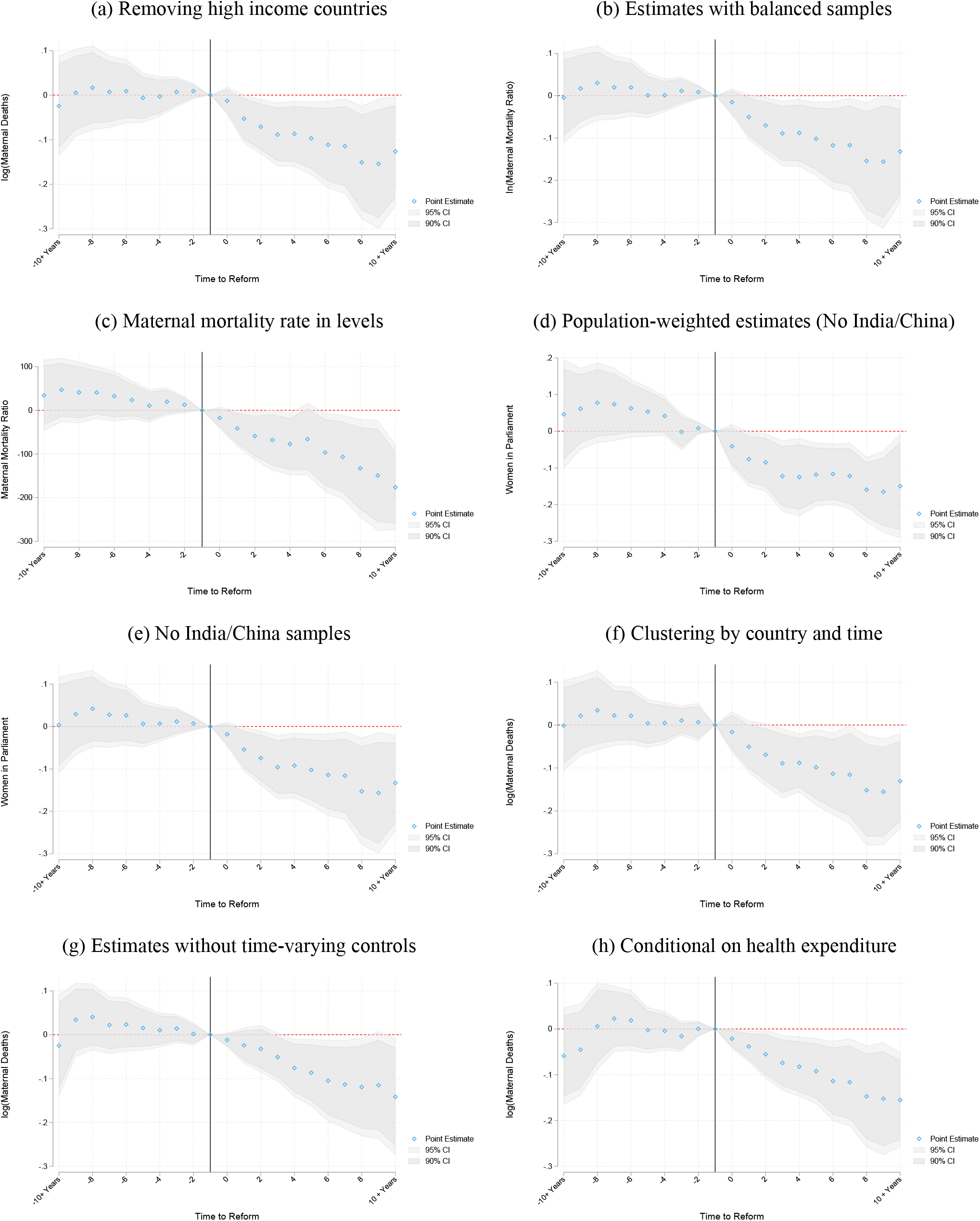
Robustness of maternal mortality reduction to alternative measures and specifications. Notes: Each panel plots event study estimates of the impact of quota passage on maternal mortality, based on alternative data sources, control groups or model specifications. Panel A removes high income countries from the control group. A static (2015) measure of high income is used to ensure consistency of the sample across years. The estimation sample of non-high-income countries consists 2,309 yearly observations in 112 countries. Panel B estimates removing any countries which legislate quotas *after* 2005, to ensure that each quota country has 10 years of lags in the event study. Panel C is presented using maternal mortality in levels rather than in logs. Panels D provides estimates using country population weights, where India and China are removed given that their population (weight) is an order of magnitude larger than most other countries. Estimates for the unweighted sample are available in panel E. Confidence intervals in Panel F are based on two-way clustering of standard errors. Panels G and H estimate with alternative specifications of controls. Identical plots for women in parliament are provided as appendix Figure B7.

A potential concern is that the availability and quality of MMR data may be endogenous if surveillance and tracking are correlated with preferences in favor of addressing MMR decline. However, any bias this creates in the coefficient of interest will render our estimates conservative– because if women parliamentarians act to expand coverage of MMR data – including remote under-developed areas for instance– then, other things equal, estimated MMR will tend to increase. Another possible concern is that inference in our specification treats the data as independent across countries, but not within countries. To address potential concerns that quota implementation was temporally correlated, we estimate event studies with two-way clustering (Cameron, Gelbach and Miller, 2011) of standard errors by both country and by year, see Figure 3f. While the confidence intervals are now wider, we still observe statistically significant effects.

#### Democracy, income and health expenditure controls

All estimates discussed so far are conditional upon income and democracy. However, because they are potentially endogenous, it is important to show that our estimates are not sensitive to inclusion of these controls (see Figure 3g and additional specifications in Figure B4). It is notable that income has no impact on the share of women in parliament, but it has a significant impact on MMR. A 1% increase in GDP results in a 0.5% reduction in MMR. A crude back-of-the-envelope calculation assuming log-linearity, and holding democracy and quotas constant suggests that to achieve the estimated 9–12% reduction in MMR due to quota adoption (on the same sample), GDP would have to increase by about 18–24%. Income is potentially endogenous so this is only a crude calculation but note that we test whether GDP changes in response to quotas and we find it does not (additional discussion is provided in section 6).

We find that democracy has direct impacts on both outcomes, increasing women in parliament and decreasing maternal mortality, but only when the democracy score is above the mean.^14^ Previous evidence on women’s sway in policy making has mostly emerged from democratic regimes, in line with theoretical models of politician behavior that admit a role for politician identity (Besley and Coate, 1997). However, a number of the quota implementing countries in our sample were non-democratic.^15^ Our results are consistent with women acting upon their innate preferences, potentially motivated by the mission of public service rather than by electoral motives.^16^

We also show that the impact of quotas on maternal mortality is robust to including health expenditure as a share of GDP as an additional control (Figure 3h). The reason that health expenditure is not included as a control in the baseline estimates is that it is only available for a subset of the sample (years 1995–2013) so controlling for it creates compositional effects.

#### Heterogeneity: Duration and dose effects, estimates by baseline MMR

We observe increasing impacts over time as displayed in Figure 2b. By 10 years out, MMR was 13% lower in countries that passed quotas. Intensive margin impacts of reserved seats that leverage variation in quota size across countries (Figure A3) are in Figure 4. The estimates are rising in quota size, consistent with a “dose-response.” The unweighted estimates from the single-coefficient DD model (Table A3) indicate that quotas of less than 15% have no significant impact, quotas of 15 to 20% raise the share of women in parliament by 5.5 percentage points and reduce MMR by 8.6% and the corresponding figures for quotas of 20–30% are 7.7 percentage points and 17.5% respectively.

**Figure 4:**
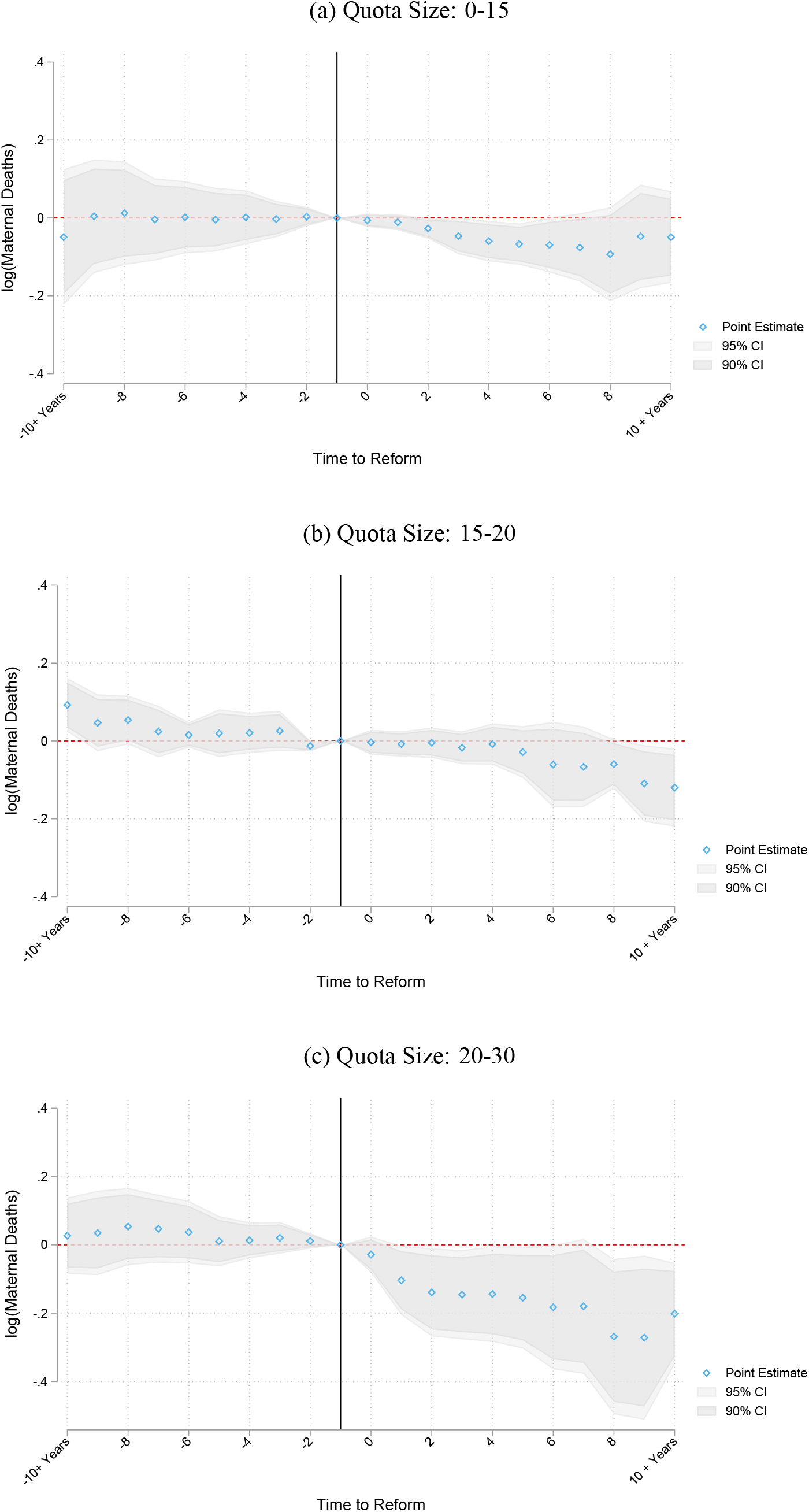
Impact of Quotas on MMR by size of gender quota. Notes: Event study estimates are presented separately for countries with a different proportion of seats reserved in their quota. These are presented in three quota size bins. Bins approximately separate quotas into three equal groups. Each set of coefficients is estimated in a single event study, and so is conditional on all other quota groups. In each case, the baseline group is countries which do not implement a quota. Remaining details follow Figure 2.

We also investigated heterogeneity in the impact of quotas by baseline MMR, dividing the sample so as to allocate roughly a third of all quota countries to each of three samples, indicated as low, medium and high rates of baseline MMR. We find a clear tendency for the impact of quotas to be higher where baseline rates are higher. Full event studies are presented in Figure 5. The summary coefficient in single term DD models is a 5.9% reduction in MMR in low baseline countries and a 18.2% reduction in high baseline countries (Table A4).

**Figure 5:**
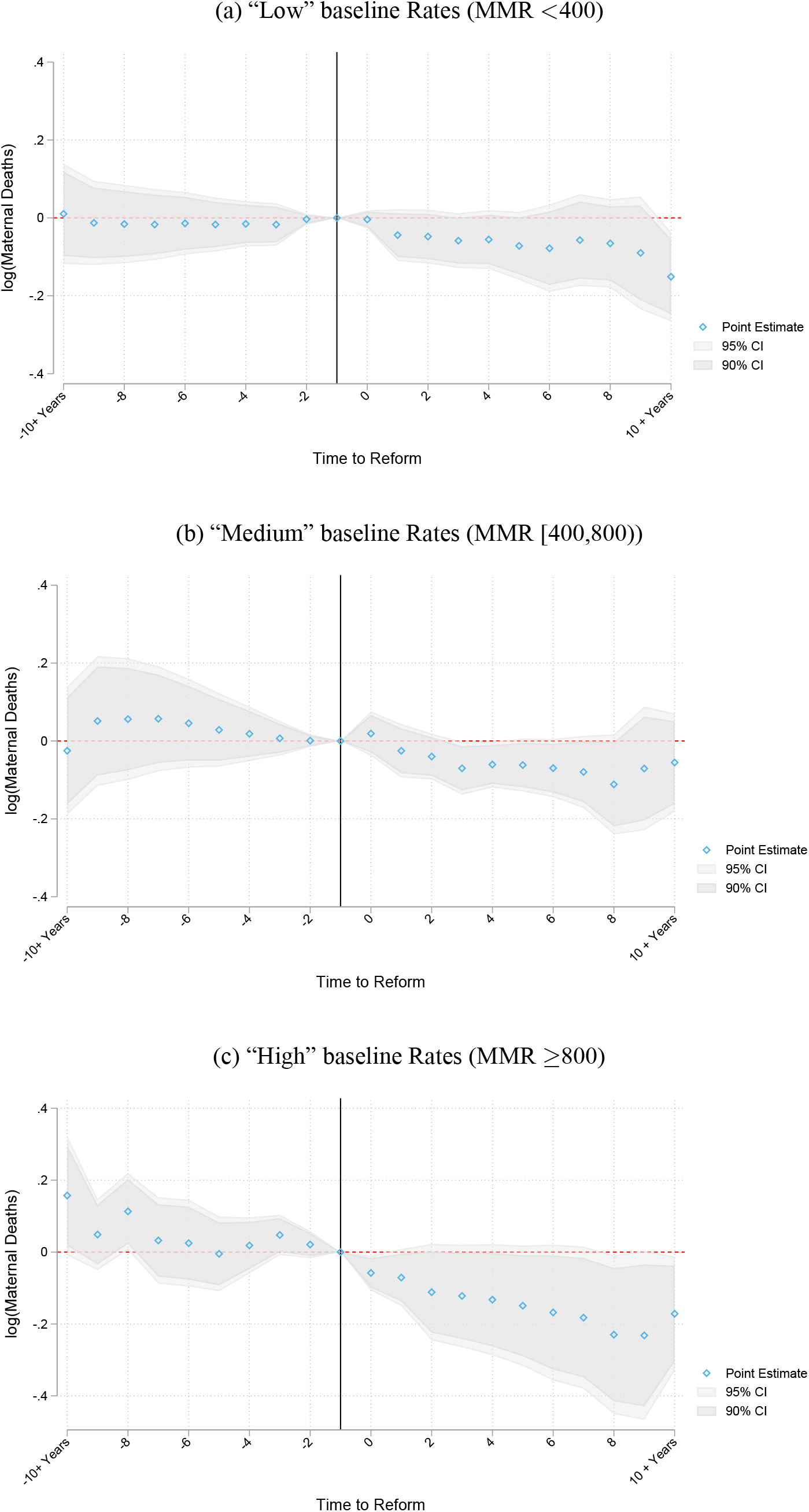
Impact of Quotas on MMR by Baseline MMR. Notes: Event study estimates are presented separately for countries with different rates of maternal mortality at baseline. Baseline maternal mortality rates are calculated as average values in countries prior to the year 2000. Ranges above are chosen to approximately equally split the number quota-passing countries into each of the three groups. Each set of lags and leads are estimated in a single model, implying that each panel is conditional on the full set quota lags/leads in each country type.

#### Omitted trends: IV and bounds

Although the event study plots mitigate potential concerns about omitted trends, we *directly* assess and address any bias in the regression coefficients associated with the possibility that when quotas were adopted, the country was already adopting other measures favorable to maternal mortality decline. To do this we estimate 2SLS regressions of MMR on the share of women in parliament, instrumented with quota implementation. Now the concern about omitted trends translates to a concern that the instrument is invalid. In particular, if quota implementation proxies a change in an omitted variable then it does not satisfy the IV exclusion restriction. However, as quota implementation is likely to be “plausibly exogenous” if not strictly so, we follow Conley, Hansen and Rossi (2012) and provide bounds on the IV estimates. The first stage is in columns 1–3 of Table A2.^17^ The second stage estimates are in Table A5. These provide the scaled impact of women’s parliamentary representation among compliers. They indicate that a 1 percentage point increase in women’s share in parliament is associated with a 1.8% decrease in MMR. In estimating bounds on the 2SLS estimates, we allow the adoption of quotas to have a direct impact on maternal mortality of up to −1% over and above its impact on MMR via increasing women in parliament. The estimated bounds are informative, indicating a 0.1% to 3.2% reduction in maternal mortality for a 1 percentage point increase in the share of women in parliament (Table A5). The unweighted estimates produce estimates in an entirely negative range, whether or not India and China are in the sample, although once we weight the bounds include zero.

#### Omitted trends: Predictors of quota legislation

As the determinants of quota legislation are of substantive interest and previous work does not provide any clear quantitative analysis, we investigated them directly by acquiring country-year panel data on predictors that have been discussed in the political science literature, typically with reference to case studies (Krook, 2010; Baines and Rubio-Marin, 2005); see Table A6. The potential predictors include evolving norms of equality and representation and accelerating movements for women’s rights, pressure from international organizations (which can be proxied with overseas development assistance), and occasions of broader constitutional reform including transitions into democracy and post-conflict reconstruction (including peace-keeping forces). Using cross-country panel data methods, we find some evidence that transitions from autocratic rulers to democracy, recent changes in women’s economic rights, and exposure to international organizations predict quota legislation (see Table A6).We include all of the potential predictors, including changes in women’s rights, as controls in the estimated equations. If the predictors of quota legislation rather than the passage of the legislation drive impacts on MMR then this would be revealed in the coefficient on quota legislation becoming insignificantly different from zero. Our estimates are, however, robust to these controls (Table A7).^18^

#### Candidate list quotas

We obtained estimates of candidate list quotas on women’s representation and found significant impacts, but smaller than associated with reserved seat quotas, a result with some antecedents (Pande and Ford, 2012; Bagues and Campa, 2017) that follows from the fact that candidate list quotas do not guarantee seats in parliament but instead require that a specified proportion of female candidates appear on the ballot.

We obtained the first estimates of impacts of candidate quotas on MMR (see Appendix Figure B5). Notice that, if we are willing to assume that the omitted variables that predict quota implementation are the same for the two sorts of quotas then this result further undermines the possibility that those omitted variables are driving our finding that reserved seat quotas lead to MMR decline.

Our finding that gender quotas on candidate lists have no discernible impact on MMR is probably a result of both (i) the smaller impact on political representation, and (ii) the fact that countries implementing candidate list quotas during the study period (predominantly in Latin America) had already achieved dramatic declines in MMR prior to quota implementation– our estimates for reserved seat quotas show that impacts are smaller where baseline rates of MMR were lower, consistent with diminishing returns to policy intervention. For most health conditions, improvements are more readily achieved when starting at higher levels.

#### Accounting for uncertainty in measurement of maternal mortality

As discussed in the Data section, the global country panel of maternal mortality data that we use were generated using a range of vital and demographic data sources (Alkema et al., 2017, 2016; World Health Organization, 2015; Wilmoth et al., 2012). We estimated the event study specification using an alternative data source providing actual (rather than estimated) maternal mortality ratios. We calculated MMR from Demographic and Health Survey (DHS) reports of maternal deaths for the 44 countries in which the DHS maternal mortality module has been implemented. We observe similar results, indicating a reduction in rates of maternal death after the implementation of quotas; see Figure 6. The timing of the decline is similar, and the effects are larger (though less precisely estimated), consistent with the DHS countries having higher MMR on average.

**Figure 6:**
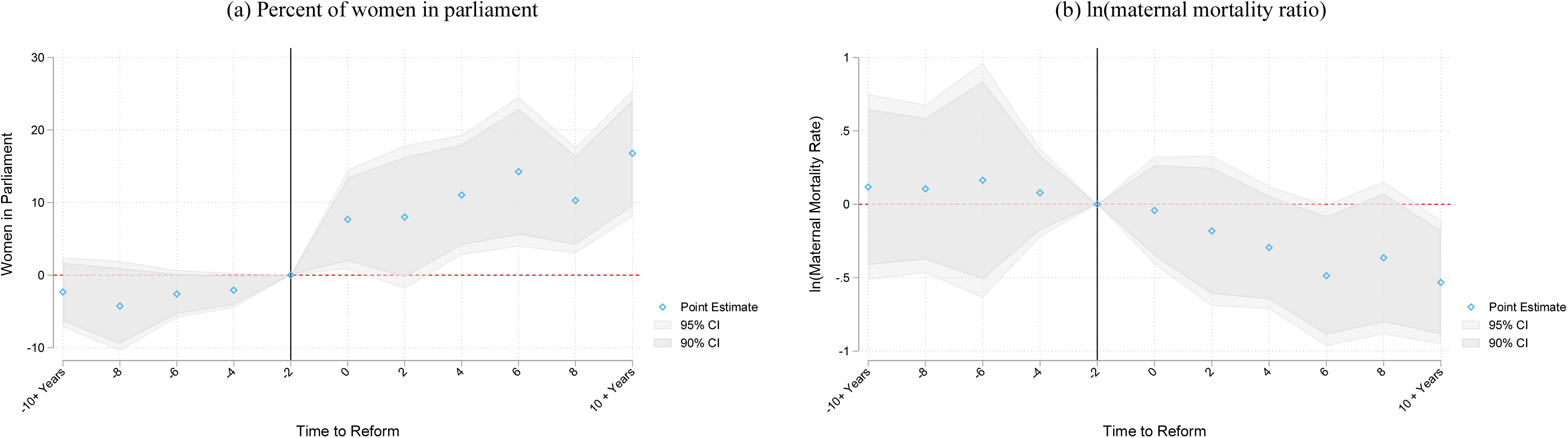
Gender quotas: Event studies from DHS microdata. Notes: Specification replicates Figure 2, however now replacing world wide estimates of MMR with maternal mortality calculated from microdata reports from the DHS. As the DHS maternal mortality module is available in only a subsample of the DHS countries (44 of 68 countries with publicly available surveys), we estimate using 2 year lags/leads to reduce noise. Substantively similar results obtain if using yearly lags and leads.

Given the multiplicity of data sources for some countries and the paucity in others, the UN estimates of MMR are modeled, and thus come with their own uncertainty intervals. We assessed if our conclusions were sensitive to directly accounting for this uncertainty. We follow a double-bootstrap procedure to calculate standard errors when undertaking inference. This procedure consists of first re-sampling observations in a (clustered) bootstrap over the countries in the original panel. We then resample the particular maternal mortality realisation for each country from within the entire uncertainty interval reported along with official maternal mortality statistics. This accounts for the (normal) sampling variation, and the uncertainty in maternal mortality data series.^19^ Appendix B describes the re-sampling algorithms and assumptions.

Table A8 replicates the difference-in-differences estimates from Table A2, showing p-values for the impact of quotas on maternal mortality associated with a range of re-sampling procedures. The different re-sampling procedures reflect different assumptions relating to the distribution of maternal mortality in the uncertainty intervals presented by the MMEIG. We first provide p-values associated with a standard clustered bootstrap prior to taking into account uncertainty in MMR measurements. Next we provide two sets of bootstrapped p-values computed assuming either a triangular distribution or a normal distribution for MMR uncertainty intervals. While the end points of the triangular distribution are at the ends of the uncertainty interval, the normal distribution provides coverage outside of the 80% UI.^20^

Table A8 shows that the triangular correction is (generally) less demanding than the normal correction, and that allowing for correlation within country reduces the estimate uncertainty. When the dependent variable is log(MMR), p-values based on the two distributional assumptions are larger than the standard bootstrapped p-values and depending on the specification fall between 0.091 and 0.226. When the dependent variable is MMR, p-values based on the two distributional assumption are larger than the standard bootstrapped p-values and fall between 0.002 and 0.069. Thus our results are robust to allowing for uncertainty in MMR estimates when the dependent variable is in levels. When it is in logarithms, robustness is more sensitive to specification.

#### Other health outcomes

In focusing on maternal mortality, we engaged the tendency for women leaders to serve the preferences of women citizens.^21^ In fact our findings are also in line with women attaching greater weight to health outcomes than men. This is plausible because women disproportionately bear the costs of bad health: women risk dying in childbirth, they bear the burden of giving birth again when children die, and they are the main caregivers for sick adults and children. Although high fertility and high morbidity characterize poor countries, the tendency for women to attach more weight to health is also evident in the British Election Survey of 2001: women expressed most concern over the quality of the National Health Service, while the single most important concern for men was low taxes (Campbell, 2004).^22^ In line with this, Miller (2008) and Bhalotra and Clots-Figueras (2014) show that women’s political participation improved infant mortality in early 19^th^ century America and contemporary India respectively.^23^

We therefore investigated whether the introduction of gender quotas led to improvements in health in other domains. For reasons discussed in section 4, we looked at adult male and female mortality and TB. See Figure B6. The event study plot for mortality among adult men suggests no significant impacts following the reform, with quite tightly estimated zeros until at least 5 years post-quota implementation, and imprecisely estimated reductions from 7 years post reform. The DD coefficient is positive but even then it is about a third of the size of the coefficient for MMR, and it collapses once we introduce population weights. For tuberculosis we similarly observe no statistically significant impact of quotas in event study specifications. The corresponding DD model yields statistically insignificant coefficients (Table B1).^24^

However if we draw a measure of infant mortality by sex from microdata in the Demographic and Health Surveys (DHS) for the sub-sample of DHS countries, and separate girls and boys, we find some evidence that gender quotas lead to lower infant mortality for girls of, on average, 12 percent (Figure 7). This is despite the fact that policies typically used to address infant mortality (e.g. provision of clean water or access to medical professionals and drugs) cannot readily discriminate between boys and girls. The reason that MMR is a conceptually clean outcome to study is that reproductive health services that address maternal mortality are by definition targeted at women.^25^

**Figure 7:**
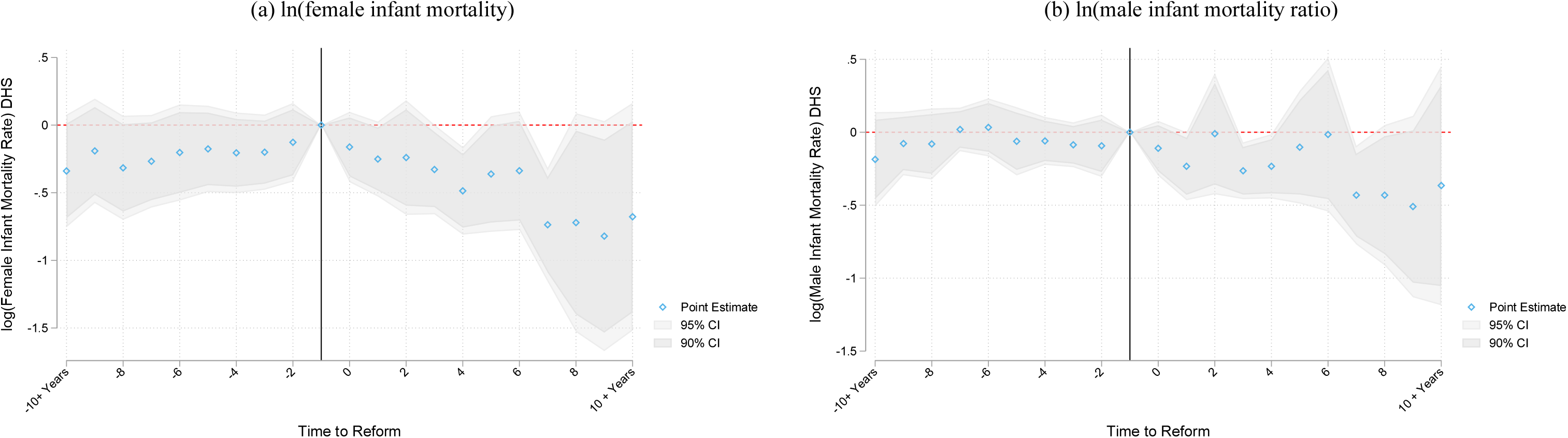
Event studies for female and male infant mortality using DHS microdata. Notes: Infant mortality is recorded from Demographic and Health Survey (DHS) microdata where mothers report their full fertility history, any children who have died, their child’s age at death (if relevant) and the year of death (if relevant). We generate infant mortality rates using retrospective fertility and survival histories for DHS surveys in the 68 publicly available DHS countries.

Overall, there are two takeaways from these results. First, women leaders appear to prioritize the health of women and girls over and above their concerns for health in general.^26^ Second, although there was no perceptible decline in male mortality or in TB mortality following gender quotas, it is notable that there was no increase as this undermines the possibility that women leaders achieved MMR decline by reallocating resources away from other areas of public health investment. Since we document below that gender quotas were not associated with increases in GDP, the share of GDP spent on public health, or international development assistance for maternal health, it seems that women effected maternal mortality decline by using available resources more effectively.

## 6 Mechanisms

Having shown that increasing the share of women in parliament leads to more rapid declines in maternal mortality, we now seek to identify underlying mechanisms. To do this, we estimated reduced form equations for endogenous mediators of the relationship between gender quotas and MMR. We consider impacts on health services interventions recommended by the WHO: antenatal care, which can help identify life threatening conditions such as pre-eclampsia and eclampsia early on; and deliveries attended by a skilled professional, which can address causes of mortality specific to the birthing process, such as uterine bleeding and post-partum infection (WHO, 2014). Simple OLS regressions of MMR on these inputs using our analysis sample and conditioning on GDP and democracy show that a 1 percentage point increase in the share of attended births or women receiving prenatal care is associated with a 3.4 and 2.3% decline in MMR respectively. These variables measure population coverage (quantity) rather than the quality of facilities (better medical equipment for existing staffed clinics), but there are no cross-country time series data indicating service quality. We also model birth rates, the education of women, health expenditure share, GDP, and international development assistance for maternal health as potential mediators. We observe that, to the extent that women parliamentarians act through information campaigns, or to improve allocative efficiency, maternal mortality decline may be achieved without any of the selected mediator variables responding to gender quotas.

We prevent event studies of these relationships, and DD specifications are provided in appendix Table B2. As discussed earlier, when the control group is large and treatment effects increase with time since event, the DD bias is likely to be small. Event study estimates of antenatal care and attended births are provided in Figure 8 panels A and B, and suggest increased rates of coverage in the years following quotas. The DD coefficients indicate a statistically significant increase of 6.9 to 9.6 percentage point in skilled birth attendance and a 5.2 to 9.9 percentage point increase in the share of women using prenatal care.

**Figure 8:**
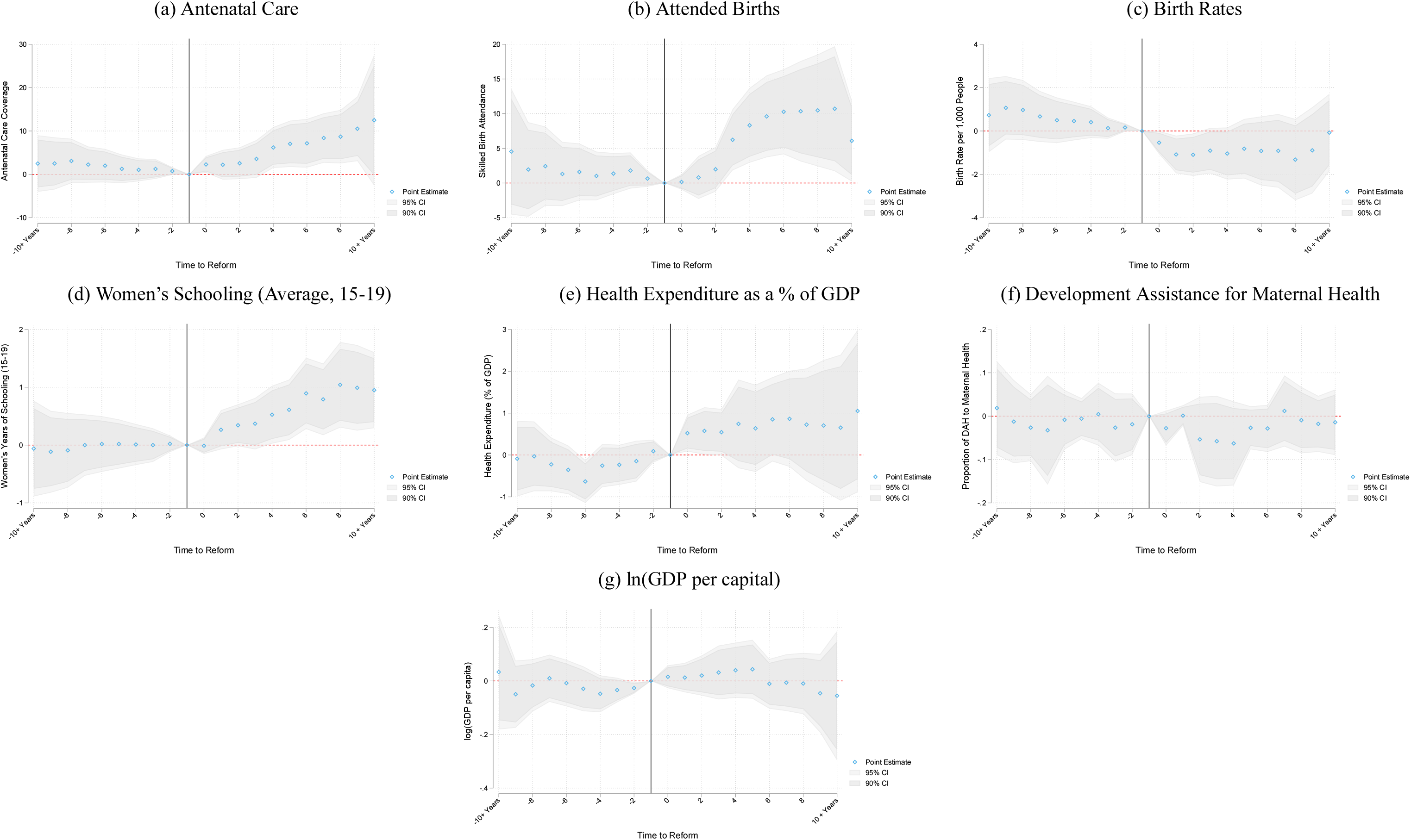
Mechanisms: Event studies for impacts of gender quotas on intermediate outcomes. Notes: Event-study estimates of intermediate outcomes as a function of the passage of gender quotas, following specification 1. Antenatal coverage and birth attendance refer to the percentage of coverage, are accessed from the World Bank databank, and are only available for a sub-sample of years for each country (an unbalanced panel from 1990–2015). Estimates in Panels A and B use a linearly interpolated measure, event studies based on non-imputed values are available in Appendix Figure B8. Birth rates are expressed as per the full population, and are available as World Bank indicator SP.DYN.CBRT.IN. Women’s schooling measures are taken from Barro-Lee which gives average years of schooling for women aged 15-19 years. Barro-Lee is only available quinquennially from 1950 to 2015. We use the sample from 1990–2015, and linearly interpolate by country between 5 year periods. Health expenditure is expressed as a percent of GDP, and is accessed from the World Health Organization’s National Health Accounts (NHA) data series. Proportion of development assistance for health that goes towards maternal health is provided by the Institute for Health Metrics and Evaluation (IHME) Development Assistance for Health Database. The log of GDP per capita is PPP adjusted and measured in 2011 international dollars.

We further investigated fertility and women’s education (Figure 8 panels C and D), given evidence that these variables are associated with maternal mortality (Bhalotra and Clarke, 2013). We find a decrease of 1.1 to 2.6 births per 1,000 population in birth rates. The DD coefficient suggests an increase in the education of girls aged 15–19 at the time of the reform by around 0.6 years, and a similar increase is observed in event studies.

We find no statistically significant increase in health expenditure as a share of GDP following introduction of quotas (Table B2), although the event study plot shows a tendency for it to increase (Figure 8e). We also find that the introduction of quotas had no significant effects on the proportion of international development assistance for health (DAH) that goes towards maternal health (see Figure 8f). This was of particular interest as DAH has played a critical role in the world’s response to the MDGs (one of which was MMR reduction) since 2000, although less so since 2010. In 2015, US$36.4 billion was disbursed in DAH, and 9.8% was for maternal health (Dieleman et al., 2016). We also tested for the possibility that gender quotas lead to higher income and that this, in turn, contributes to MMR decline. We see no evidence of any trend break in GDP following the passage of quotas (Figure 8g).

Women leaders could have effected declines in MMR at low cost by disseminating information, campaigning, monitoring, or inspiring action (Miller, 2008; Baskaran et al., 2018; Beaman et al., 2009; Bhalotra and Clots-Figueras, 2014).^27^

## 7 Conclusion

We show that women’s involvement in policy-making can effect rapid maternal mortality decline and at low cost. Thus gender quotas may be a powerful at-scale means of modifying public health priorities in favour of maternal health. Despite significant progress, especially since 2000, preventable maternal mortality remains high. The lifetime risk of maternal mortality is 1 in 41 women in low income countries. Despite a wave of gender quota implementation, 130 countries in the world have none. There is thus substantial room for manoeuvre. Our findings have implications for the recently launched Global Health 2035 report, and the ambitious Sustainable Development Goals. The MDG target was not met and the SDG target is more ambitious, so we clearly need some policy innovation. In fact this paper shows that SDG 3.1 for reducing maternal mortality is complementary to SDG 5.5 for raising the share of women in parliament.

## Data Availability

All data and code for this article will be posted on a public repository when the article is published. We are happy to provide any interested parties with the code and dataset in advance with a written request.

## Appendix Figures and Tables

**Figure A1:**
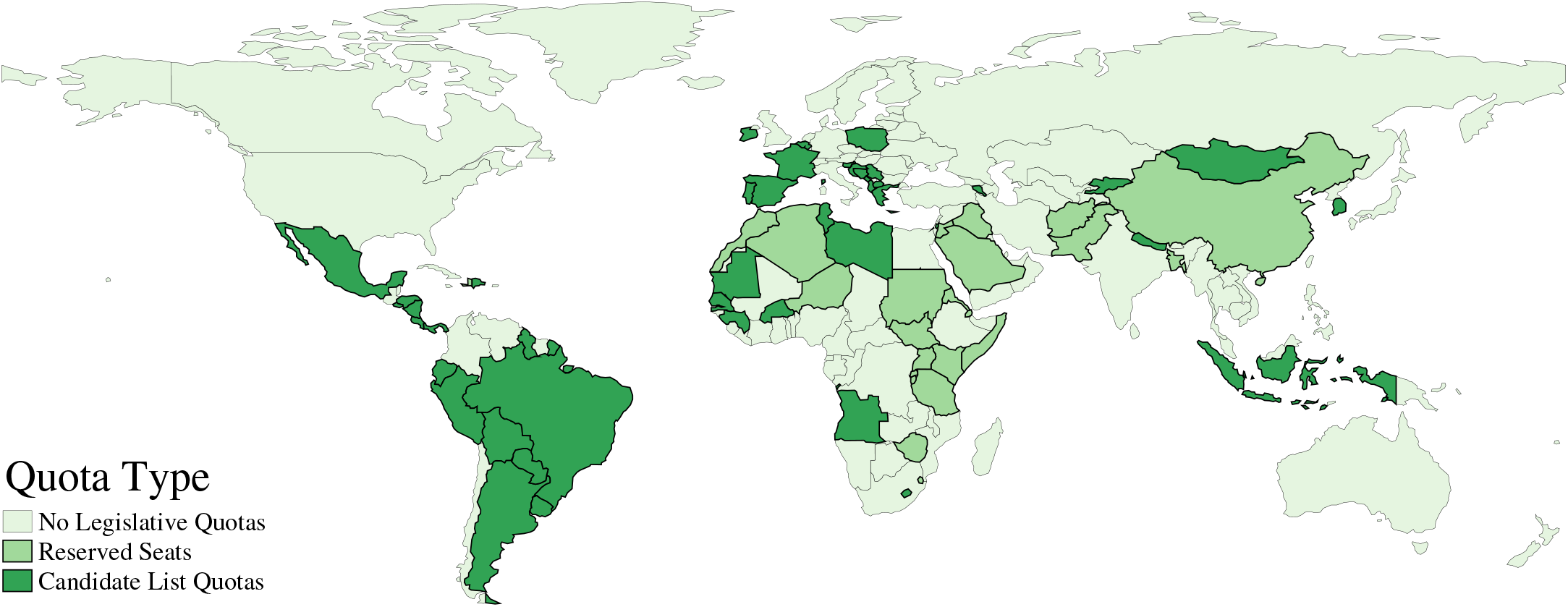
Quota coverage: 1990–2015. Notes: Geographic distribution of countries implementing a quota for reserved seats in parliament and candidate list quotas. Data compiled from Dahlerup (2005) and updated with information for recent years from the online quotaproject.org database developed and maintained by the International Institute for Democracy and Electoral Assistance (IDEA), the Inter-Parliamentary Union, and Stockholm University. This database was consulted on 19th of July, 2016.

**Figure A2:**
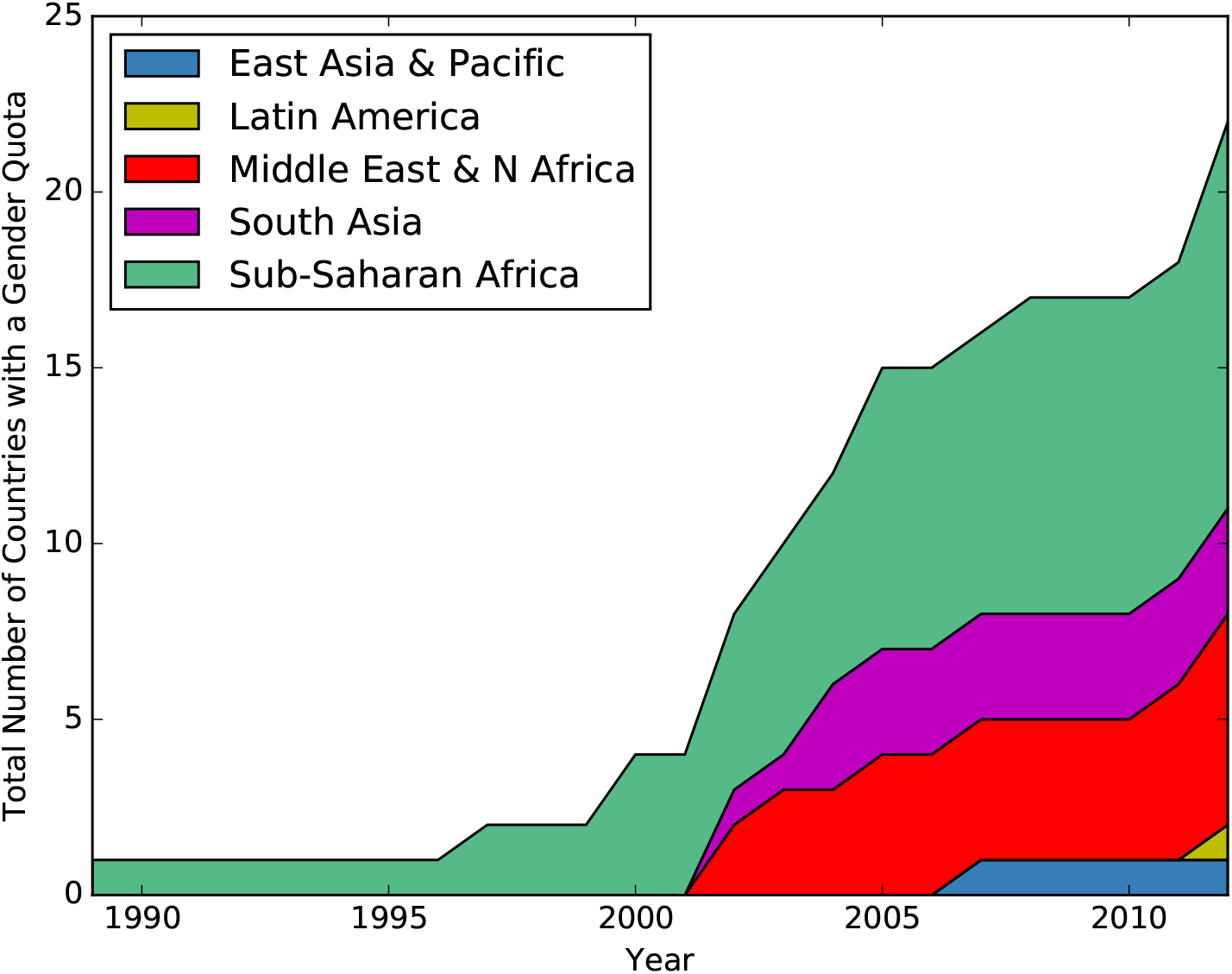
Reserved seat quota timing: 1990–2012. Notes: Timing of the implementation of reserved seats by geographic area. Additional notes in Figure A1.

**Figure A3:**
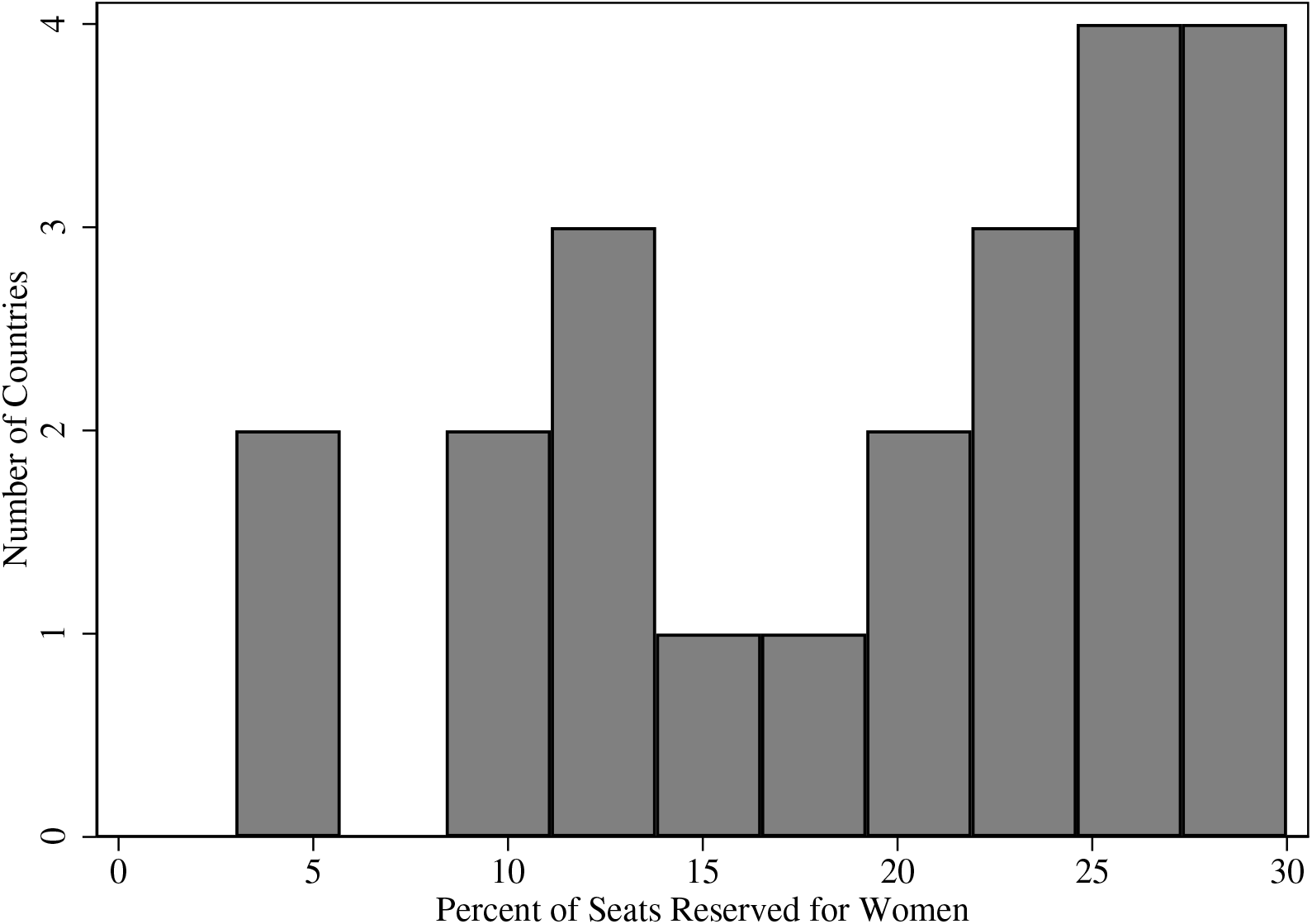
Reserved seat quota sizes. Notes: This histogram describes the quota size for each country which adopts a reserved seat quota. Each country (quota) is included as a single observation.

**Figure A4:**
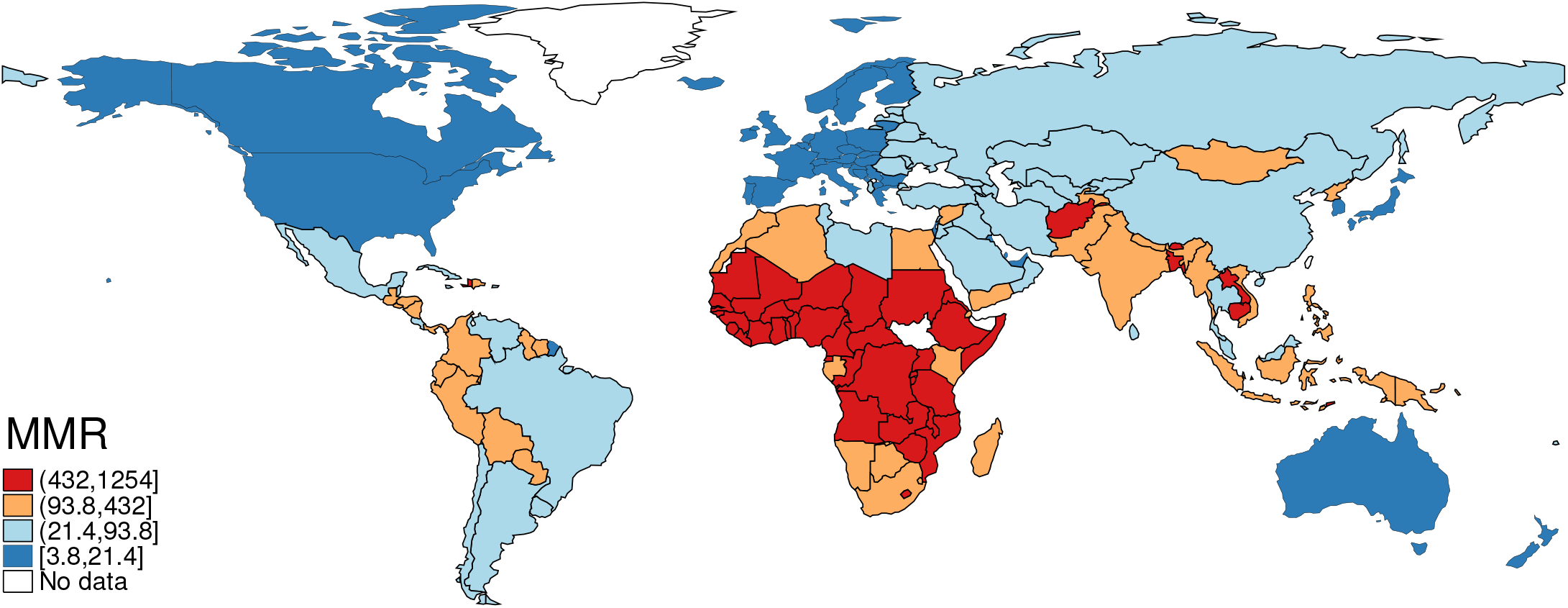
Maternal mortality ratio: 1990–2015. Notes: Average rates by country for the period 1990–2015. Values are calculated as deaths per 100,000 live births, and are provided by WHO, UNICEF, UNFPA, World Bank Group, and the United Nations Population Division.

**Table A1:** Summary statistics for reserved seat analysis

|  | N | Mean | Std. Dev. | Min | Max |
| --- | --- | --- | --- | --- | --- |
| % Women in Parliament | 4186 | 14.06 | 10.45 | 0.00 | 63.80 |
| Maternal Mortality Ratio | 4186 | 236.03 | 325.90 | 3.00 | 2890.00 |
| Reserved Seats | 4186 | 0.06 | 0.24 | 0.00 | 1.00 |
| Male Mortality Rate (15-60) | 4126 | 240.90 | 120.48 | 58.80 | 753.70 |
| Female Mortality Rate (15-60) | 4126 | 168.08 | 116.62 | 34.32 | 685.03 |
| ln(GDP per capita) | 4186 | 8.90 | 1.22 | 5.51 | 11.77 |
| Polity IV Democracy score | 3212 | 5.60 | 3.85 | 0.00 | 10.00 |
| Percent of Pregnancies Receiving Prenatal Care | 662 | 84.04 | 17.75 | 15.40 | 100.00 |
| Percent of Births Attended by Skilled Staff | 1199 | 83.33 | 24.16 | 5.00 | 100.00 |
| Health Expenditure as a % of GDP | 3124 | 6.24 | 2.39 | 0.72 | 17.10 |
| Female Infant Mortality Rate (DHS subsample) | 1067 | 0.08 | 0.05 | 0.00 | 0.60 |
| Male Infant Mortality Rate (DHS subsample) | 1066 | 0.10 | 0.05 | 0.00 | 0.33 |
| Birth rates per 1,000 population | 4160 | 24.30 | 11.74 | 7.60 | 55.56 |

Notes: Refer to Data Appendix A for a full description of each variable and its source. The Maternal Mortality Ratio is measured as deaths per 100,000 live births. For comparison, the male and female mortality rates for 15–60 year-olds is expressed as per 1,000 male and female adults respectively. Reserved seats is a binary variable taking one for each country and year pair where a quota was implemented, and 0 otherwise.

**Table A2:** Gender quotas: DD impacts on women in parliament and maternal mortality

|  | % Women in Parliament |  |  | ln(Maternal Mortality Ratio) |  |  | Maternal Mortality Ratio |  |  |
| --- | --- | --- | --- | --- | --- | --- | --- | --- | --- |
|  | (1) | (2) | (3) | (4) | (5) | (6) | (7) | (8) | (9) |
| Reserved Seats | 5.144**<br>[2.301] | 5.818**<br>[2.326] | 6.051***<br>[1.860] | -0.107**<br>[0.049] | -0.113**<br>[0.051] | -0.148**<br>[0.062] | -110.599**<br>[49.926] | -124.659**<br>[50.661] | -115.805***<br>[24.891] |
| Mean of Dep. Var. | 13.647 | 13.632 | 13.632 | 4.397 | 4.389 | 4.389 | 249.190 | 249.707 | 249.707 |
| Observations | 3212 | 3167 | 3167 | 3212 | 3167 | 3167 | 3212 | 3167 | 3167 |
| Number of Countries | 156 | 154 | 154 | 156 | 154 | 154 | 156 | 154 | 154 |
| R-Squared | 0.475 | 0.483 | 0.565 | 0.597 | 0.588 | 0.621 | 0.375 | 0.382 | 0.454 |
| Population Weights | N | N | Y | N | N | Y | N | N | Y |

Difference-in-differences estimates of the impact of reserved seats in parliament on women in parliament (columns 1-3), the log of the maternal mortality ratio (columns 4-6), and MMR in levels (columns 7-9). In each case country and year fixed effects are included, and time-varying controls consist of the log of PPP adjusted GDP per capita, and a democracy score. Unweighted (columns 1-2, 4-5, and 7-8), and population weighted specifications (column 3, 6 and 9) are displayed. When weighting, China and India are removed from the estimation sample, to avoid regression results being largely driven by these two countries, with a population an order of magnitude larger than other countries. The unweighted specification without these countries is displayed in columns 2, 5 and 8. Standard errors clustered by country are displayed in parentheses. \* $p < 0.10$ ; \*\* $p < 0.05$ ; \*\*\* $p < 0.01$ .

**Figure A5:**
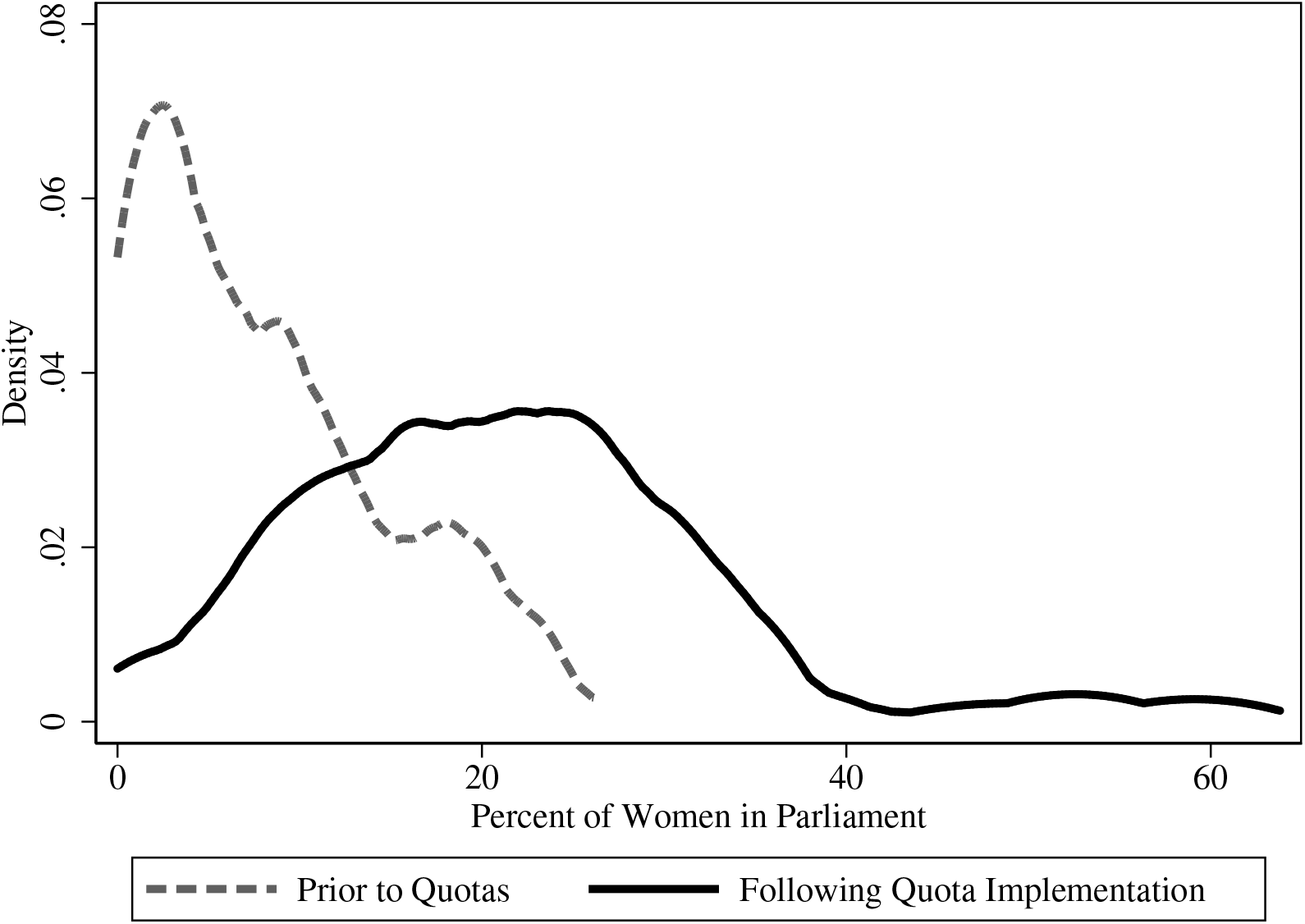
Proportion of women in parliament in countries with reserved seats. Notes: Density plots for the proportion of women in parliament in countries which at some point adopt a reserved seat quota. Plots are based on each country by year observation in the women in parliament data.

**Table A3:**
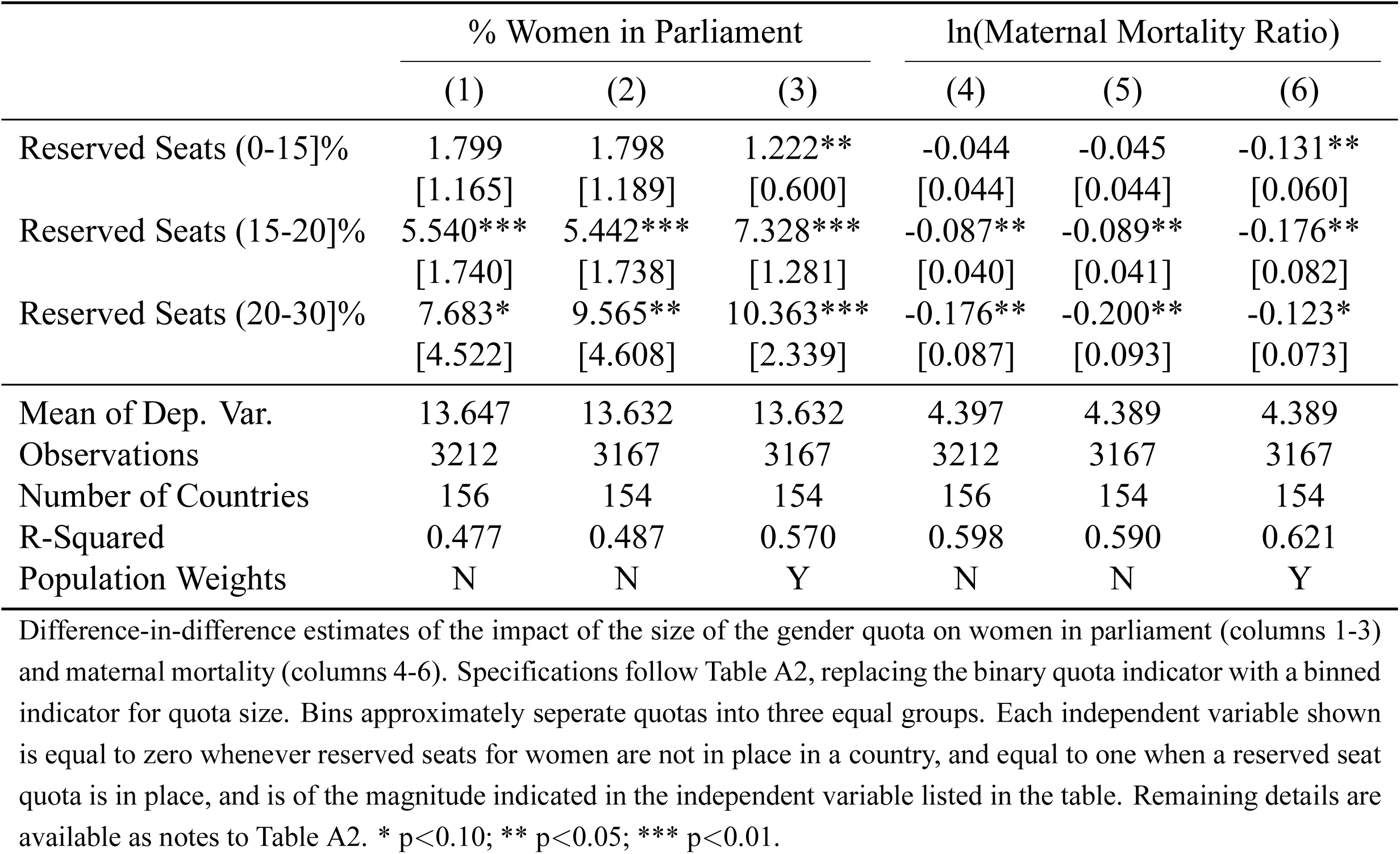
Intensive margin impacts of reserved seats (binned by quota size)

|  | % Women in Parliament |  |  | ln(Maternal Mortality Ratio) |  |  |
| --- | --- | --- | --- | --- | --- | --- |
|  | (1) | (2) | (3) | (4) | (5) | (6) |
| Reserved Seats (0-15]% | 1.799<br>[1.165] | 1.798<br>[1.189] | 1.222**<br>[0.600] | -0.044<br>[0.044] | -0.045<br>[0.044] | -0.131**<br>[0.060] |
| Reserved Seats (15-20]% | 5.540***<br>[1.740] | 5.442***<br>[1.738] | 7.328***<br>[1.281] | -0.087**<br>[0.040] | -0.089**<br>[0.041] | -0.176**<br>[0.082] |
| Reserved Seats (20-30]% | 7.683*<br>[4.522] | 9.565**<br>[4.608] | 10.363***<br>[2.339] | -0.176**<br>[0.087] | -0.200**<br>[0.093] | -0.123*<br>[0.073] |
| Mean of Dep. Var. | 13.647 | 13.632 | 13.632 | 4.397 | 4.389 | 4.389 |
| Observations | 3212 | 3167 | 3167 | 3212 | 3167 | 3167 |
| Number of Countries | 156 | 154 | 154 | 156 | 154 | 154 |
| R-Squared | 0.477 | 0.487 | 0.570 | 0.598 | 0.590 | 0.621 |
| Population Weights | N | N | Y | N | N | Y |

Difference-in-difference estimates of the impact of the size of the gender quota on women in parliament (columns 1-3) and maternal mortality (columns 4-6). Specifications follow Table A2, replacing the binary quota indicator with a binned indicator for quota size. Bins approximately separate quotas into three equal groups. Each independent variable shown is equal to zero whenever reserved seats for women are not in place in a country, and equal to one when a reserved seat quota is in place, and is of the magnitude indicated in the independent variable listed in the table. Remaining details are available as notes to Table A2. \* $p < 0.10$ ; \*\* $p < 0.05$ ; \*\*\* $p < 0.01$ .

**Table A4:** Impacts of reserved seats by baseline MMR

|  | % Women in Parliament |  |  | ln(Maternal Mortality Ratio) |  |  |
| --- | --- | --- | --- | --- | --- | --- |
|  | (1) | (2) | (3) | (4) | (5) | (6) |
| Reserved Seats (Low Baseline MMR) | 2.540<br>[2.360] | 4.311**<br>[2.063] | 8.271***<br>[1.492] | -0.059*<br>[0.034] | -0.072**<br>[0.033] | -0.181**<br>[0.089] |
| Reserved Seats (Mid Baseline MMR) | 3.595**<br>[1.631] | 3.556**<br>[1.644] | 2.070**<br>[0.893] | -0.066<br>[0.056] | -0.067<br>[0.056] | -0.125**<br>[0.057] |
| Reserved Seats (High Baseline MMR) | 9.038*<br>[5.345] | 9.030*<br>[5.310] | 10.146***<br>[3.216] | -0.182*<br>[0.108] | -0.183*<br>[0.109] | -0.142<br>[0.087] |
| Mean of Dep. Var. | 13.647 | 13.632 | 13.632 | 4.397 | 4.389 | 4.389 |
| Observations | 3212 | 3167 | 3167 | 3212 | 3167 | 3167 |
| Number of Countries | 156 | 154 | 154 | 156 | 154 | 154 |
| R-Squared | 0.480 | 0.487 | 0.574 | 0.598 | 0.589 | 0.622 |
| Population Weights | N | N | Y | N | N | Y |

Difference-in-difference estimates of the impact of the size of the gender quota on women in parliament (columns 1-3) and maternal mortality (columns 4-6). Specifications follow Table A2, replacing the binary quota indicator with a series of quota indicators, indicating if a country adported a reserved seat quota, and if its baseline maternal mortality was in the group ( $<400$ ), the group (400-800) or the group ( $\geq 800$ ). Magnitudes approximately separete quotas into three equal groups. Each independent variable shown is equal to zero whenever reserved seats for women are not in place in a country, and equal to one when a reserved seat quota is in place, and baseline MMR is of the magnitude described. Remaining details are available as notes to Table A2. \* $p < 0.10$ ; \*\* $p < 0.05$ ; \*\*\* $p < 0.01$ .

**Table A5:** Reserved seats as an IV for women in parliament

|  | (1) | (2) | (3) |
| --- | --- | --- | --- |
|  | ln(MMR) | ln(MMR) | ln(MMR) |
| % Women in Parliament | -0.020***<br>[0.007] | -0.019***<br>[0.006] | -0.025**<br>[0.012] |
| F-Statistic First Stage | 4.753 | 5.951 | 10.070 |
| p-value First Stage | 0.031 | 0.016 | 0.002 |
| 95% CI from Conley et al. (2012) | [-0.035;-0.005] | [-0.032;-0.005] | [-0.049;0.049] |
| Mean of Dep. Var. | 4.397 | 4.389 | 4.389 |
| Observations | 3212 | 3167 | 3167 |
| Number of Countries | 156 | 154 | 154 |
| Population Weights | N | N | Y |

Instrumental variables regressions are run where gender quotas are used to instrument women in parliament. The first stage regression of women in parliament on reserved seats is displayed in columns 1-3 of Table A2. F-Statistic of the first stage and the associated p-value are traditional tests of instrumental relevance. Displayed coefficients give the effect of an additional percentage of women in parliament on rates of maternal mortality, where women in parliament is instrumented with reserved seats. The 95% confidence interval from Conley et al. (2012) is a robustness test, where we allow the instrument to be imperfect in the sense that the exclusion restriction is only close to holding. These confidence intervals are associated with the estimates where quotas are able to have a direct effect in reducing MMR that is *not* mediated by women in parliament of 0.01 (or 1%) using Conley et al. (2012)'s Union of Confidence Intervals (UCI) approach. Each regression includes country and year fixed effects and clusters standard errors by country. \* p<0.10; \*\* p<0.05; \*\*\* p<0.01.

**Table A6:** The passage of reserved seat legislation

|  | No Country Fixed Effects |  |  | Country Fixed Effects |  |  |
| --- | --- | --- | --- | --- | --- | --- |
|  | (1) | (2) | (3) | (4) | (5) | (6) |
| Overseas Development Assistance | 0.003<br>[0.018] | -0.008<br>[0.020] | -0.023<br>[0.028] | -0.021<br>[0.022] | -0.018<br>[0.029] | -0.032<br>[0.033] |
| Peace Keepers | 0.002<br>[0.002] | 0.016**<br>[0.008] | 0.018*<br>[0.010] | 0.003**<br>[0.001] | 0.017**<br>[0.008] | 0.020**<br>[0.010] |
| Change in Women's Rights | 0.008*<br>[0.005] | 0.008<br>[0.005] | 0.009*<br>[0.005] | 0.008*<br>[0.005] | 0.008<br>[0.005] | 0.009*<br>[0.005] |
| Right Wing Executive | -0.002<br>[0.002] | -0.002<br>[0.003] | -0.002<br>[0.003] | -0.002<br>[0.002] | -0.000<br>[0.003] | -0.000<br>[0.003] |
| Left Wing Executive | -0.003<br>[0.003] | -0.003<br>[0.003] | -0.003<br>[0.003] | -0.004<br>[0.004] | -0.004<br>[0.005] | -0.003<br>[0.005] |
| Years in Power | -0.000<br>[0.000] | -0.000*<br>[0.000] | -0.000<br>[0.000] | -0.001***<br>[0.000] | -0.001***<br>[0.000] | -0.001***<br>[0.000] |
| Herfindahl Index | -0.010<br>[0.007] | -0.012<br>[0.007] | -0.012<br>[0.008] | -0.014<br>[0.011] | -0.017<br>[0.012] | -0.018<br>[0.014] |
| Vote Share Opposition | -0.000<br>[0.000] | -0.000<br>[0.000] | -0.000<br>[0.000] | -0.000<br>[0.000] | -0.000<br>[0.000] | -0.000<br>[0.000] |
| Transitioning Regime | 0.008<br>[0.005] | 0.009<br>[0.006] | 0.009<br>[0.007] | 0.009<br>[0.006] | 0.011<br>[0.007] | 0.012<br>[0.008] |
| First Lag (ODA) |  | 0.030<br>[0.029] | 0.011<br>[0.030] |  | 0.010<br>[0.029] | -0.001<br>[0.029] |
| First Lag (peace keepers) |  | -0.015**<br>[0.008] | -0.021<br>[0.015] |  | -0.015*<br>[0.008] | -0.022<br>[0.015] |
| First Lag ($\Delta$ Womens Rights) | | -0.003<br>[0.004] | -0.001<br>[0.003] | | -0.003<br>[0.004] | -0.001<br>[0.003] |
| Second Lag (ODA) |  |  | 0.036<br>[0.030] |  |  | 0.017<br>[0.025] |
| Second Lag (peace keepers) |  |  | 0.004<br>[0.007] |  |  | 0.006<br>[0.008] |
| Second Lag ($\Delta$ Womens Rights) | | | 0.005<br>[0.005] | | | 0.005<br>[0.005] |
| Observations | 1951 | 1840 | 1729 | 1951 | 1840 | 1729 |
| Number of Countries | 118 | 117 | 117 | 118 | 117 | 117 |
| R-Squared | 0.023 | 0.044 | 0.047 | 0.027 | 0.045 | 0.050 |

Each column regresses a variable indicating whether a quota law was passed in a given year on potential predictors of quota adoption suggested in the political science literature. Each specification includes year fixed effects and standard errors are clustered by country. Overseas Development Assistance (ODA) measured as net inflows in current US dollars divided by GDP in current US dollars is generated from the World Bank Data Bank. Peacekeepers (measured in 1000s) are from the IPI Peacekeeping Database, changes in women's rights refer to changes in economic rights for women as compiled by the CIRI Human Rights Data Project, and political measures including the orientation of leader's party, the time in power, Herfindahl Index of parties, vote shares and regime types and changes are recorded by the Database of Political Institutions. Additional lags of relevant variables are included in columns 2 and 3, and 5 and 6. \* $p < 0.10$ ; \*\* $p < 0.05$ ; \*\*\* $p < 0.01$ .

**Table A7:** Estimates including all potential quota predictors

|  | ln(Maternal Mortality Ratio) |  | % Women in Parliament |  |
| --- | --- | --- | --- | --- |
|  | (1) | (2) | (3) | (4) |
| Reserved Seats | -0.096* | -0.109** | 5.419** | 6.465** |
|  | [0.051] | [0.052] | [2.353] | [2.876] |
| Overseas Development Assistance |  | 0.042 |  | -4.288 |
|  |  | [0.073] |  | [4.375] |
| Peace Keepers |  | -0.002 |  | 0.052 |
|  |  | [0.002] |  | [0.147] |
| Change in Women's Rights |  | -0.001 |  | 0.257 |
|  |  | [0.006] |  | [0.254] |
| Right Wing Executive |  | 0.015 |  | -0.934 |
|  |  | [0.038] |  | [0.696] |
| Left Wing Executive |  | -0.064 |  | 1.265 |
|  |  | [0.060] |  | [0.833] |
| Years in Power |  | -0.001 |  | 0.095** |
|  |  | [0.001] |  | [0.042] |
| Herfindahl Index |  | -0.062 |  | 1.318 |
|  |  | [0.053] |  | [1.348] |
| Vote Share Opposition |  | -0.000 |  | -0.020 |
|  |  | [0.000] |  | [0.013] |
| Transitioning Regime |  | -0.023* |  | 0.801 |
|  |  | [0.013] |  | [0.506] |
| First Lag (ODA) |  | 0.019 |  | -1.698 |
|  |  | [0.048] |  | [2.342] |
| Second Lag (ODA) |  | -0.021 |  | 0.169 |
|  |  | [0.075] |  | [2.432] |
| First Lag (peace keepers) |  | -0.000 |  | -0.081 |
|  |  | [0.003] |  | [0.207] |
| Second Lag (peace keepers) |  | -0.001 |  | 0.018 |
|  |  | [0.004] |  | [0.178] |
| First Lag ($\Delta$ Womens Rights) | | 0.003 | | 0.223 |
|  |  | [0.008] |  | [0.292] |
| Second Lag ($\Delta$ Womens Rights) | | -0.002 | | 0.152 |
|  |  | [0.006] |  | [0.206] |
| Observations | 2310 | 1641 | 2310 | 1641 |
| Number of Countries | 113 | 109 | 113 | 109 |
| R-Squared | 0.610 | 0.618 | 0.444 | 0.479 |
| Proposed Predictors | N | Y | N | Y |

The regressions include country and year fixed effects and controls for log GDP and a democracy index. All potential predictors of quotas, described in Table A6, are included as controls. \* $p < 0.10$ ; \*\* $p < 0.05$ ; \*\*\* $p < 0.01$ .

**Table A8:** Alternative Inference Procedures for Measures of Maternal Mortality in Principal Diff-in-Diff Specification

|  | ln(Maternal Mortality Ratio) |  |  | Maternal Mortality Ratio |  |  |
| --- | --- | --- | --- | --- | --- | --- |
|  | (1) | (2) | (3) | (4) | (5) | (6) |
| Reserved Seats (Point Estimate) | -0.088 | -0.093 | -0.120 | -102.169 | -114.949 | -106.907 |
| p-value Bootstrap | 0.096 | 0.090 | 0.115 | 0.040 | 0.020 | 0.002 |
| p-value Triangular Correction | 0.117 | 0.112 | 0.149 | 0.044 | 0.022 | 0.002 |
| p-value Triangular Correction by Country | 0.098 | 0.091 | 0.118 | 0.046 | 0.024 | 0.002 |
| p-value Normal Correction | 0.200 | 0.190 | 0.226 | 0.055 | 0.030 | 0.007 |
| p-value Normal Correction by Country | 0.097 | 0.092 | 0.113 | 0.069 | 0.039 | 0.005 |
| Mean of Dep. Var. | 4.397 | 4.389 | 4.389 | 249.190 | 249.707 | 249.707 |
| Observations | 3212 | 3167 | 3167 | 3212 | 3167 | 3167 |
| Number of Countries | 156 | 154 | 154 | 156 | 154 | 154 |
| R-Squared | 0.596 | 0.587 | 0.619 | 0.373 | 0.378 | 0.449 |
| Population Weights | N | N | Y | N | N | Y |

Difference-in-differences estimates of the impact of reserved seats in parliament on log of the maternal mortality ratio (columns 1-3), and MMR in levels (columns 4-6) replicate results from Table A2 of the working paper. Point estimates are presented, and below p-values associated with each point estimate, based on different procedures for re-sampling the uncertainty of measures of maternal mortality. In each case, re-samples are taken over country clusters, as treatment is defined at the level of the country.

### Online Appendix – Not for Print

**Figure B1:**
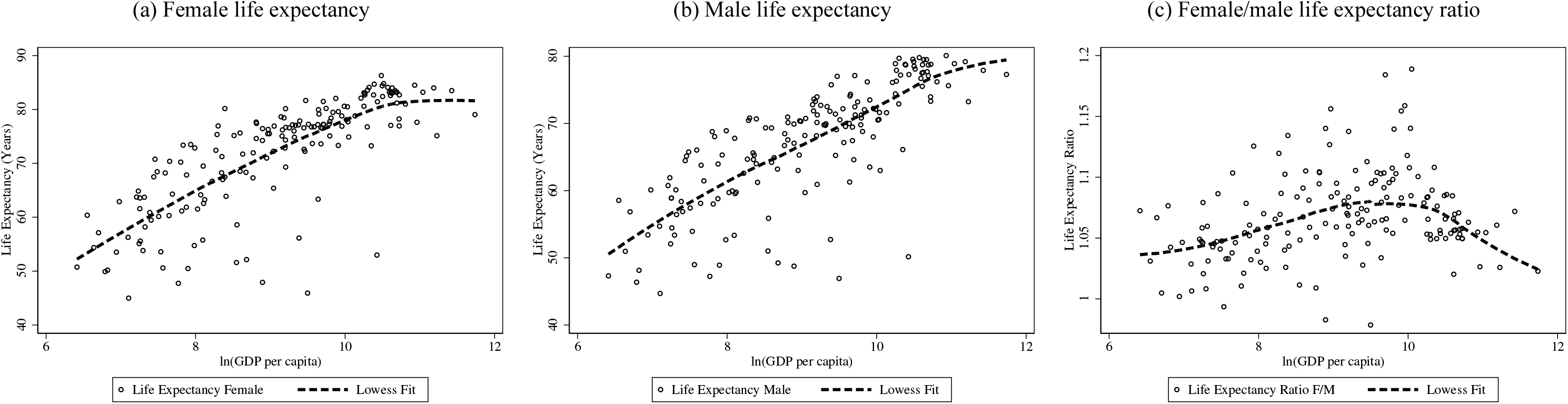
Life expectancy in 2010. Notes: Life expectancy at birth and PPP-adjusted GDP per capita data are collated by the World Bank Data Bank. These indicators are SP.DYN.LE00.FE.IN (Female life expectancy) SP.DYN.LE00.MA.IN (Male life expectancy) and NY.GDP.PCAP.PP.KD (PPP adjusted GDP per capita). The life expectancy ratio is calculated as female life expectancy divided by male life expectancy for each country. Lowess fits are overlaid on scatter plots, using a bandwidth of 0.8 for local linear smoothing.

**Figure B2:**
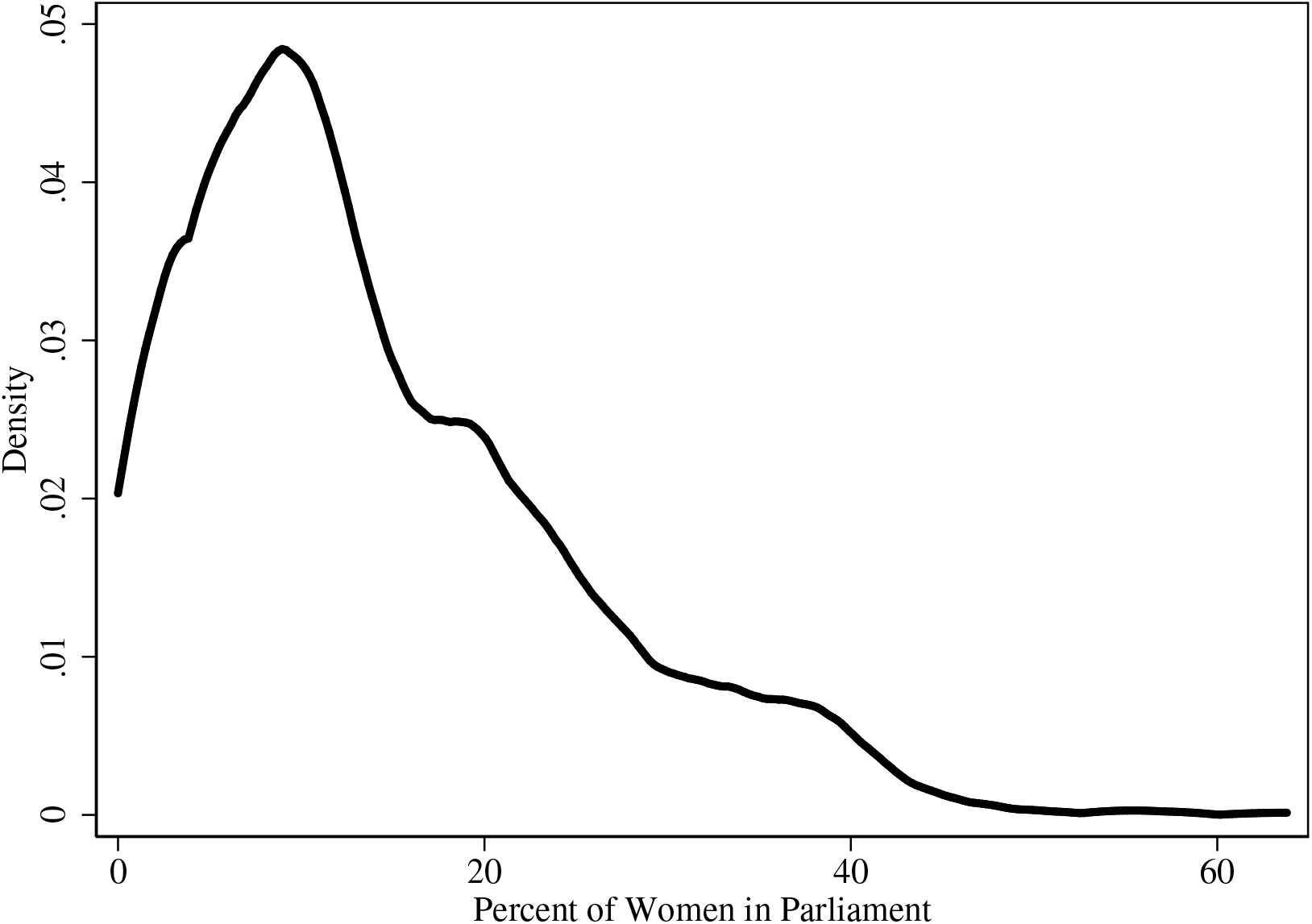
Proportion of women in parliament in all countries. Notes: Density plots describe the proportion of women in parliament in all countries and years under study.

**Figure B3:**
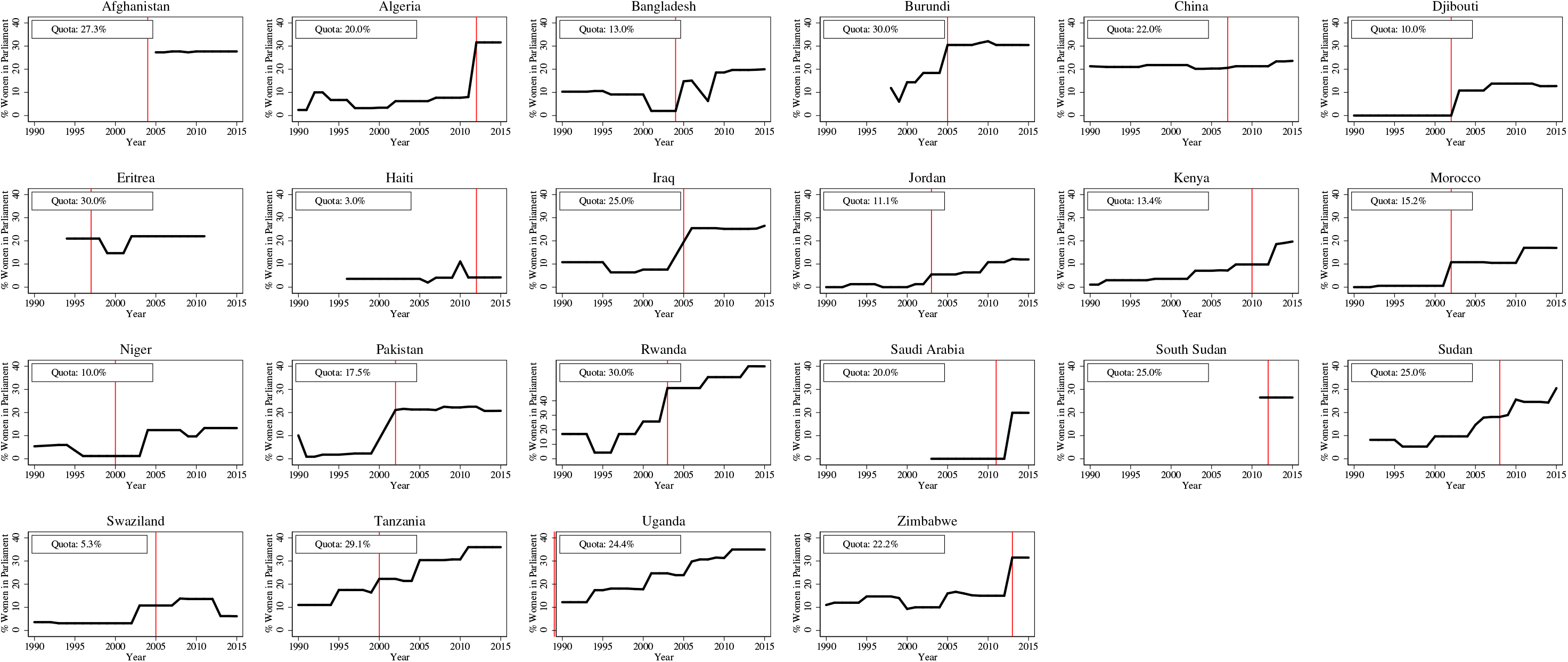
Country-specific changes in women in parliament after reserved seat quotas. Notes: In each panel, the vertical lines display the recorded date of the passage of a reserved seat quota for women in the national parliament, and the plots show the evolution of the percentage of women in parliament. Each figure shares a common y-axis for ease of comparison.

**Figure B4:**
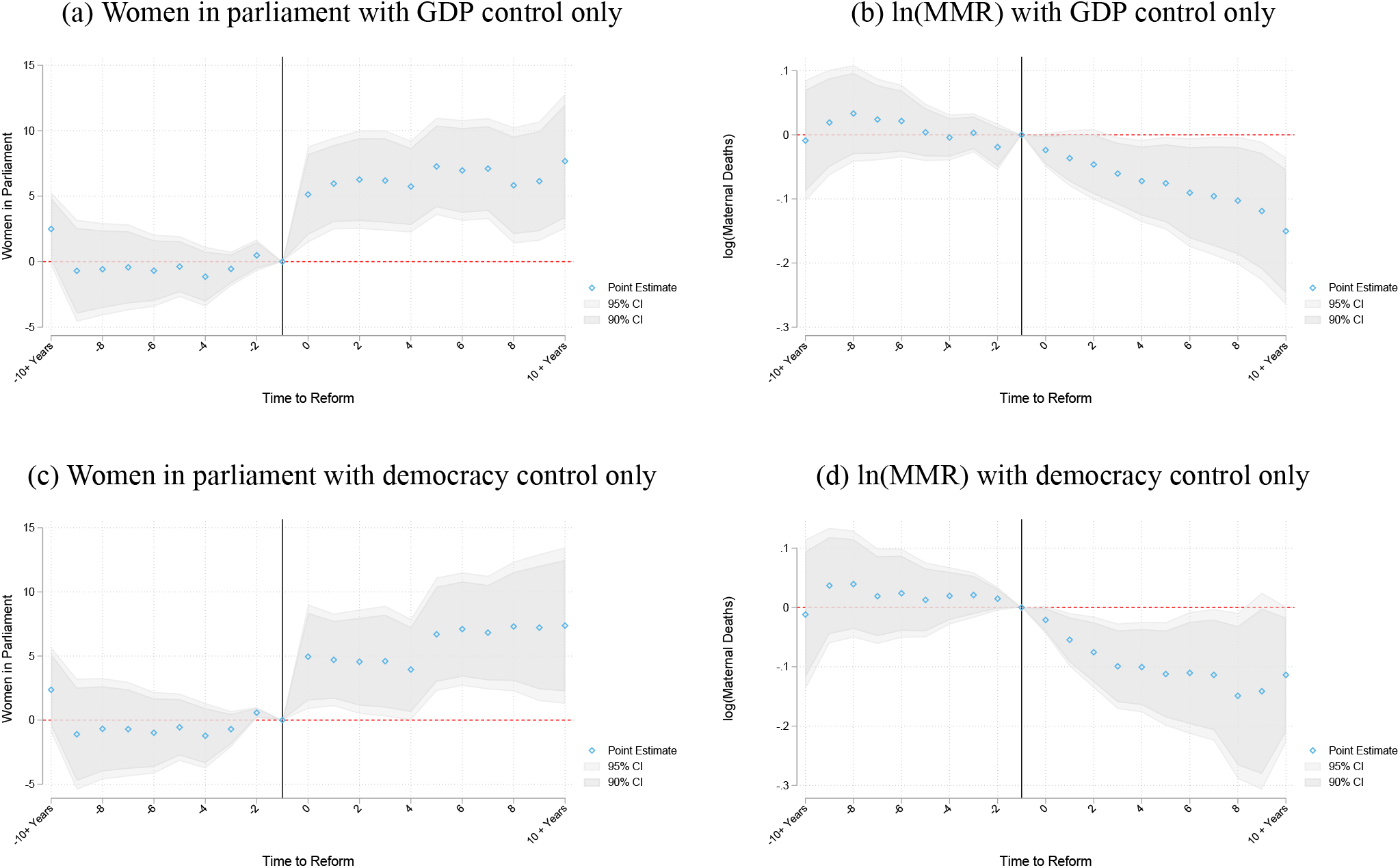
Alternative specification of quota event study. Notes: Alternative specifications of the event study shown in Figure 2. Specifications are shown with and without included controls, and with only GDP or only democracy controls. Results are robust to population weights, and additionally controlling for health spending per capita. Additional notes in Figure 2.

**Figure B5:**
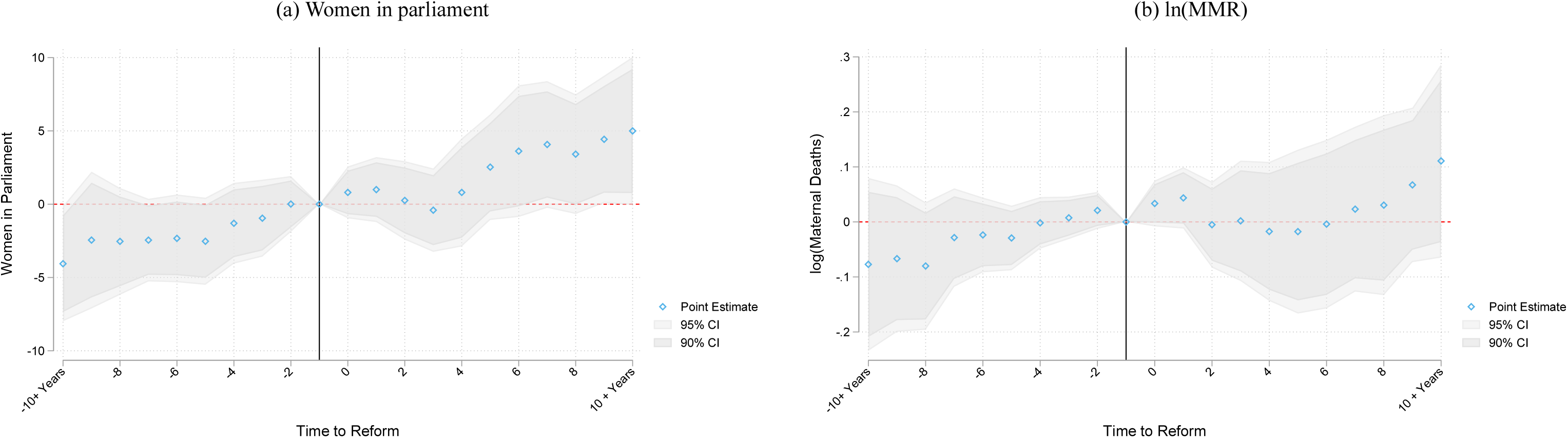
Candidate List Quotas: Event studies for women in parliament and maternal mortality. Notes: Identical specifications are estimated as in Figure 2, however now with countries passing candidate list quotas. Difference-in-difference estimates from a single-coefficient model suggest an increase in 1.776 (standard error 2.172) in the proportion of women in parliament following reserved seat quotas, and a reduction of 0.017 (standard error 0.071) in log(MMR). Analogous values for unweighted estimates are 3.569 (s.e. 1.180) for women in parliament, and 0.032 (s.e. 0.055) for log(MMR). Countries implementing candidate list quotas in the period under study are Albania, Angola, Argentina, Armenia, Belgium, Bolivia, Bosnia and Herzegovina, Brazil, Burkina Faso, Costa Rica, Croatia, Dominican Republic, Ecuador, El Salvador, France, Greece, Guinea, Guyana, Honduras, Indonesia, Ireland, South Korea, Kyrgyz Republic, Lesotho, Macedonia, Mauritania, Mexico, Mongolia, Montenegro, Nepal, Nicaragua, Panama, Paraguay, Peru, Poland, Portugal, Senegal, Serbia, Slovenia, Spain, Tunisia and Uruguay.

**Figure B6:**
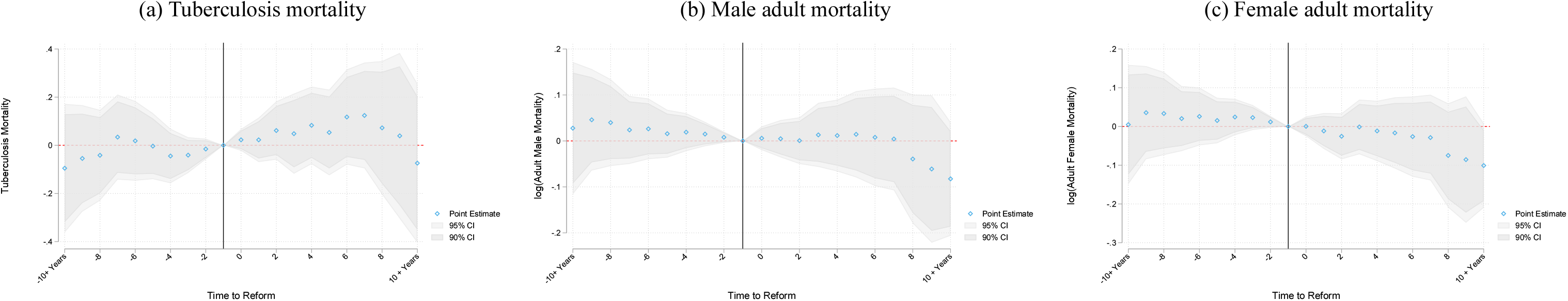
Event Studies for the impact of gender quotas on TB (gender neutral) and adult male and female mortality. Notes: Identical event studies are plotted to those in Figure 2 examining quota impacts on alternative health outcomes. These outcomes are the rate of death due to TB per 100,000 individuals, and infant mortality per 1,000 live births, male mortality per 1,000 adult males (ages 15–60), and female mortality per 1,000 adult females (ages 15–60). For comparison with Figure 2b, the natural logarithm of each variable is used, with the exception of TB mortality, where an inverse sine transformation is used, given a small number of country-year observations where a rate of zero is observed.

**Table B1:** Gender quotas: DD impacts on TB mortality and Adult Mortality

|  | ln(TB Mortality) |  |  | ln(Male Mortality) |  |  | ln(Female Mortality) |  |  |
| --- | --- | --- | --- | --- | --- | --- | --- | --- | --- |
|  | (1) | (2) | (3) | (4) | (5) | (6) | (7) | (8) | (9) |
| Reserved Seats | 0.084<br>[0.121] | 0.110<br>[0.124] | -0.018<br>[0.090] | -0.027<br>[0.058] | -0.027<br>[0.061] | -0.010<br>[0.059] | -0.047<br>[0.056] | -0.050<br>[0.059] | -0.075*<br>[0.045] |
| Mean of Dep. Var. | 2.691 | 2.680 | 2.680 | 5.391 | 5.394 | 5.394 | 4.925 | 4.926 | 4.926 |
| Observations | 3212 | 3167 | 3167 | 3208 | 3163 | 3163 | 3208 | 3163 | 3163 |
| Number of Countries | 156 | 154 | 154 | 156 | 154 | 154 | 156 | 154 | 154 |
| R-Squared | 0.409 | 0.403 | 0.544 | 0.496 | 0.491 | 0.588 | 0.490 | 0.484 | 0.620 |
| Population Weights | N | N | Y | N | N | Y | N | N | Y |

Difference in difference specification following that in Table A2, with identical controls and fixed effects. We now examine estimates for impacts of gender quotas on tuberculosis and male and female mortality rates. Tuberculosis is measured as incidence per 100,000 people, and mortality is measured as per 1,000 adults of ages 15-60. Further notes are available in Table A2. Standard errors clustered by country are displayed in parentheses. \* $p < 0.10$ ; \*\* $p < 0.05$ ; \*\*\* $p < 0.01$ .

**Figure B7:**
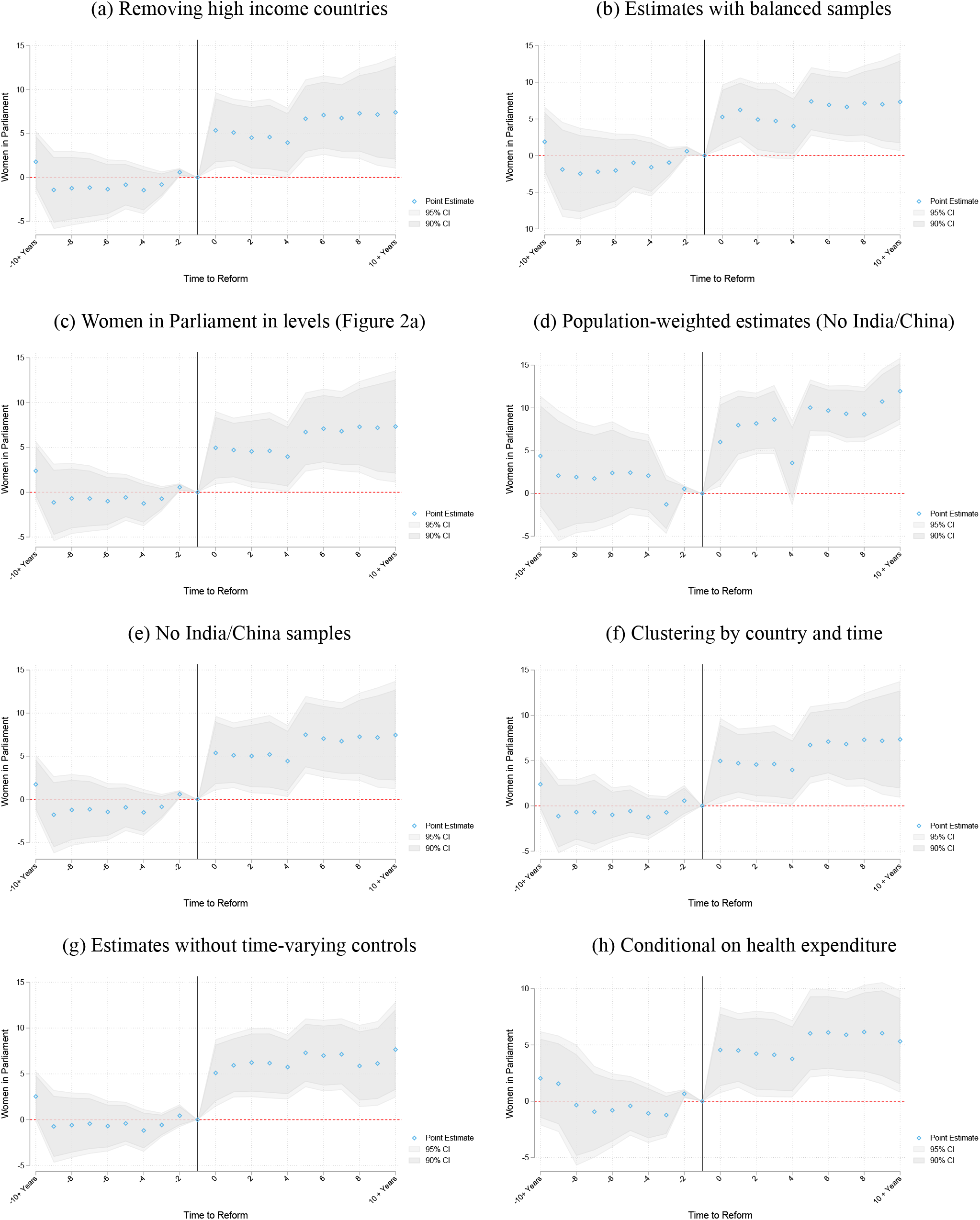
Robustness of impact of quotas on women in parliament to alternative measures and specifications. Notes: Plots replicates Figure 3 of the paper, however now showing the impact of quotas on women in parliament. Notes available in Figure 3.

**Figure B8:**
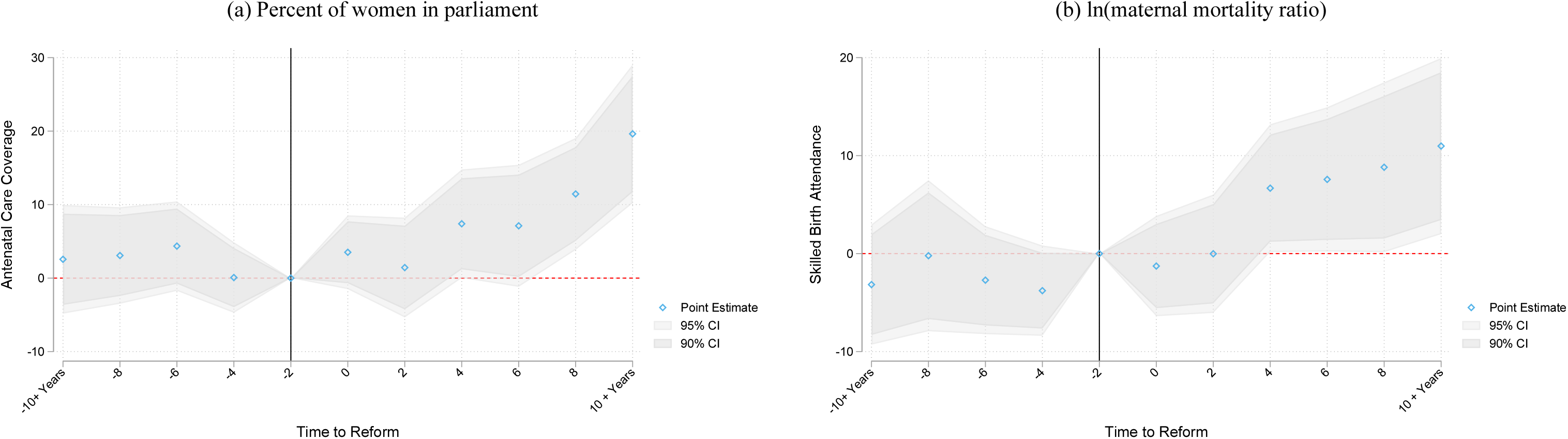
Mechanisms: Event studies for impacts of quotas on non-interpolated antenatal care and birth attendance. Notes: Event studies replicate panels A and B of Figure 8 using non-interpolated data. Given the unbalanced coverage of antenatal care and attended birth measures by countries and years, we present estimates for these outcomes pooling in 2 yearly bins, rather than yearly bins, to avoid unbalanced coverage in particular lag and lead terms where possible.

**Table B2:** Mechanisms: impacts of gender quotas on intermediate outcomes

|  | Antenatal Care |  |  | Attended Births |  |  | Birth Rate |  |  |
| --- | --- | --- | --- | --- | --- | --- | --- | --- | --- |
|  | (1) | (2) | (3) | (4) | (5) | (6) | (7) | (8) | (9) |
| Reserved Seats | 5.168<br>[3.544] | 8.229**<br>[3.151] | 9.884***<br>[2.737] | 6.932**<br>[3.236] | 9.597***<br>[2.849] | 8.214***<br>[2.222] | -1.121<br>[0.731] | -1.162<br>[0.770] | -2.642***<br>[0.949] |
| Mean of Dep. Var. | 82.673 | 82.641 | 82.641 | 83.024 | 82.964 | 82.964 | 24.489 | 24.549 | 24.549 |
| Observations | 526 | 500 | 500 | 996 | 970 | 970 | 3189 | 3144 | 3144 |
| Number of Countries | 134 | 132 | 132 | 149 | 147 | 147 | 155 | 153 | 153 |
| R-Squared | 0.507 | 0.524 | 0.725 | 0.362 | 0.373 | 0.689 | 0.584 | 0.579 | 0.605 |
| Population Weights | N | N | Y | N | N | Y | N | N | Y |

Difference in difference models of intermediate outcomes as a function of the passage of gender quotas, where gender quotas is the second lag of the passage of quota laws. In each case, we control for the log of PPP adjusted GDP per capita, and a democracy score. Antenatal care coverage and birth attendance are newly harmonized data available for 1990-2015, however only available in a sub-sample of years for each particular country. Birth rates are per 1,000 inhabitants. Unweighted, and population weighted specifications are displayed. When weighting, China and India are removed from the estimation sample, to avoid regression results being largely driven by these two countries with a population an order of magnitude larger than other countries. \* $p < 0.10$ ; \*\* $p < 0.05$ ; \*\*\* $p < 0.01$ .

**Table B3:** Mechanisms: impacts of gender quotas on intermediate outcomes

|  | Years of Schooling |  |  | Health Expenditure |  |  | Development Assistance |  |  | ln(GDP per capita) |  |  |
| --- | --- | --- | --- | --- | --- | --- | --- | --- | --- | --- | --- | --- |
|  | (1) | (2) | (3) | (4) | (5) | (6) | (7) | (8) | (9) | (10) | (11) | (12) |
| Reserved Seats | 0.588**<br>[0.259] | 0.593**<br>[0.272] | 1.321***<br>[0.413] | 0.735<br>[0.478] | 0.740<br>[0.511] | -0.209<br>[0.344] | -0.024<br>[0.026] | -0.026<br>[0.027] | -0.085**<br>[0.034] | 0.012<br>[0.058] | -0.024<br>[0.046] | 0.044<br>[0.052] |
| Mean of Dep. Var. | 7.462 | 7.464 | 7.464 | 6.132 | 6.157 | 6.157 | 0.095 | 0.095 | 0.095 | 8.899 | 8.910 | 8.910 |
| Observations | 2480 | 2439 | 2439 | 2586 | 2550 | 2550 | 2445 | 2400 | 2400 | 3212 | 3167 | 3167 |
| Number of Countries | 132 | 130 | 130 | 155 | 153 | 153 | 127 | 125 | 125 | 156 | 154 | 154 |
| R-Squared | 0.357 | 0.346 | 0.441 | 0.229 | 0.230 | 0.451 | 0.126 | 0.125 | 0.231 | 0.458 | 0.453 | 0.668 |
| Population Weights | N | N | Y | N | N | Y | N | N | Y | N | N | Y |

### A Data Appendix

#### Maternal Mortality Data

We used recently released estimates of the maternal mortality ratio (MMR) per 100,000 live births produced by the Maternal Mortality Estimation Inter-Agency Group (MMEIG) and published in the World Bank World Development Indicators (WDI, indicator SH.STA.MMRT). These data were made available for the first time in the year 2016 and before that there were no reliable annual cross-country data on MMR. These estimates were available for 183 countries annually for the period 1990–2015. Maternal mortality is identified using ICD-10 codes O00-O99 (Pregnancy, childbirth and puerperium); the official definition is “the number of women who die from pregnancy-related causes while pregnant or within 42 days of pregnancy termination per 100,000 live births.” These are widely considered the best MMR measures to date, as they address known measurement difficulties in survey and vital statistics data on maternal mortality using Bayesian methods applied to multiple, complementary data sources including vital statistics, special inquiries, surveillance sites, population-based household surveys and census files (Alkema et al., 2016, 2017). The world distribution of average MMR for the period of 1990–2015 is in Figure A4.

#### Political Gender Quota Data

We collated measures for each country of whether the country has a legislated and binding reserved seat quota for women, its year of implementation, and the size of the quota measured as number of seats divided by all seats in the uni- or bi-cameral chamber. To create the database, we started with measures provided by Dahlerup (2005) and completed the most recent years from Global Database of Quotas for Women database (available online at quotaproject.org), which is a repository developed and maintained by the International Institute for Democracy and Electoral Assistance (IDEA), the Inter-Parliamentary Union, and Stockholm University.

#### Women in Parliament Data

We used three distinct annual-level measures of women in parliament to construct a comprehensive panel of the percentage of women occupying seats in the national parliament. These were the WDI indicator SG.GEN.PARL.ZS (“Proportion of seats held by women in national parliaments (%)”), The UN Millennium Development Goals (MDG) Indicators (“Seats held by women in national parliament, percentage”), and the Interuniversity Consortium for Political and Social Research (ICPSR) dataset compiled by (Paxton, Green and Hughes, 2008) (“Women in Parliament, 1945–2003: Cross-National Dataset”). The first two of these datasets had partially-complete coverage for the years 1990, and then 1997–2015, while the latter had partially-complete yearly coverage for each year starting in 1945, and ending in 2003. In order to construct as comprehensive a series as possible, we began with the WDI data, and then imputed missing years where available from the MDG indicators, and Paxton, Green and Hughes (2008) data. When a missing WDI year was available in both the MDG and the ICPSR dataset, we favored the MDG measure, which was estimated using the same sample and year. Figures A5 and B2 present the distribution of the proportion of women in parliament pre- and post-quota implementation in quota countries, as well as the full distribution of the proportion of women in parliament over the period under study.

#### Covariates

We adjusted for the natural logarithm of PPP adjusted GDP per capita measured in 2011 international dollars, and a score for the level of democracy in the country, in all models. In additional sensitivity tests, we also examined quota predictors as laid out in Krook (2010). These were the number of peacekeepers in a country from The International Peace Institute, IPI Peacekeeping Database, Net Overseas Development Assistance (World Bank Indicator DT.ODA.ODAT.CD), and a series of measures of political competition and landscape from (Beck et al., 2001). Our measure of democracy was gleaned from the Polity IV project database. This database records information on the political regime in 167 countries, between 1800 and 2014. The democracy indicator is available annually, and is a 0–10 scale based on measures of competitiveness of political participation, openness and competitiveness of executive recruitment and constraints on executive powers. Higher values reflect more open, democratic societies.

Health expenditure at the country-year level was taken from the World Health Organization the National Health Accounts (NHA) data series. These provide a measure of total health expenditure as a percent of GDP, and are available for the years 1995-2013.

The data on development assistance for health are based on the Institute for Health Metrics and Evaluation (IHME) Development Assistance for Health Database (1990–2017). These data are available at the source country × receiver country × year level. We compute the proportion Development Assistance for Health to Maternal Health as: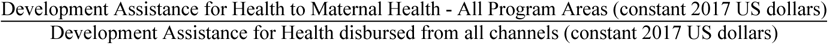.

For the women’s economic rights variable we exploit a previously under-exploited cross-country rights data from the Cingranelli, Richards and Clay (2013) data set, which provides data on three different variables measuring Political, Economic and Social Rights of women, for the period of 1981 to 2011 for around 127 (in 1981) to 192 (in 2011) countries.

#### Maternal Care Inputs Data

Recent data from the World Bank Data Bank allow us to examine the state of maternal health care in a sub-set of countries and years. We use the two policy-relevant indicators measuring the percent of pregnant women receiving prenatal care (indicator SH.STA.ANVC.ZS) and the percent of all births attended by skilled health staff (indicator SH.STA.BRTC.ZS). These data are constructed and released by the World Bank using comparable measures from each country: specifically data from UNICEF, the State of the World’s Children, ChildInfo, and the Demographic and Health Surveys. As such, these measures are only available in years and countries for which surveys were conducted, resulting in fewer observations than the yearly measures of maternal mortality. In our analysis we use the full set of data released in the World Bank Data Bank.

#### Placebo Outcomes

Data on male mortality for adults are available in the World Bank Data Bank (indicator SP.DYN.AMRT.MA), based on measures from the United Nations Population Division, World Population Prospect and University of California, Berkeley, and Max Planck Institute for Demographic Research. This is measured as mortality between the ages of 15–60, per 1,000 male adults, and captures the likelihood that a male of age 15 dies by the age of 60. Tuberculosis mortality is measured as the number of deaths due to Tuberculosis among HIV-negative people, and is measured per 100,000 population. The data are from the WHO and were downloaded from: http://apps.who.int/gho/data/view.main.57020ALL?lang=en, accessed on 17/03/2016.

### B Description of Resampling Procedure for MMR Uncertainty

Publicly available MMR data consist of a point estimate and the upper and lower points of the 80% uncertainty interval. In order to estimate standard errors and p-values based on these data, we undertake the following procedure:

#### Resample Algorithm

1. Take a clustered bootstrap resample from the original data *b* = 1, …, *B*, with *B* = 500
2. Generate a random vector of size *N* (where *N* is the sample in the regression) where each element is either a) a draw of a normal variable where 80% of the probability mass falls between −1 and 1 (a draw from 𝒩 (0, 0.7803)), or b) a draw from a triangular distribution in the interval [-1,1]. Below this is ***ε***, and in each case, these integrate to 1.
3. Generate a resampled value of maternal mortality as: 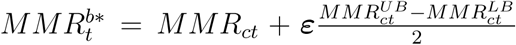, where 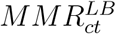 is the lower bound estimate and 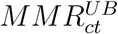 is the upper bound estimate; ie take the original measure, and draw a value from the uncertainty interval centred around this measure.
4. Estimate the original regression using the resampled data, with the re-resampled MMR measure. This results in an estimate of interest 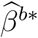
5. if *b <* 500 return to step 1. Else go to step 6
6. Calculate the standard error of 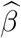 as the standard deviation of 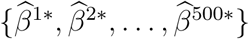. This replaces the original naive standard error, and similarly a p-value can be calculated associated with the null hypothesis of a null impact, analogous to the p-value calculated based on a standard regression coefficient.

This procedure is only necessary when estimating impacts of quotas on maternal mortality, and not for women in parliament as we are only adjusting for uncertainty in the dependent variable in cases where maternal mortality is used. Note that in the above we are re-sampling maternal mortality to provide full coverage of the 80% uncertainty interval, or indeed, to provide greater than full coverage in the case of the normal draw, for each country year pair. In each case, the normal or triangular distribution places more weight on the likelihood of observing a value of maternal mortality close to the stated estimate, and less weight on the likelihood of observing a value in the tails of the distribution.

The above resampling procedure assumes that uncertainty in maternal mortality is independent between countries and years. It may also be the case that uncertainty is correlated across years within a country. When undertaking inference robust to uncertainty we thus present p-values associated with a range of cases as presented in Table A8. These are:

1. Bootstrap: The bootstrap analogue of the original p-value (ie no uncertainty in MMR)
2. Triangular Correction: Resamples from the MMR uncertainty range from the WHO data (80% coverage) with a triangular distribution whose minimum and maximum are at the end points of the uncertainty range, and whose centre is at the estimate
3. Triangular Correction by Country: Resamples as above, however now instead of taking uncertainty draws by country and year, takes uncertainty draws only at the level of the country. This implies that uncertainty with regards to MMR is perfectly correlated within a country over time. It is the limit case of assuming correlation within a country in uncertainty in MMR measurement.
4. Normal Correction: Resamples from the MMR uncertainty range assuming a normal distribution, where draws are taken so that the 10^th^/90^th^ quintile of the normal are at the upper and lower end points of the uncertainty range presented in the WHO data in each case. This allows for us to sample outside the 80% confidence bounds presented in the original data, and will be the most demanding of all corrections.
5. Normal Correction by Country: Resamples as above, however now instead of taking uncertainty draws by country and year, takes uncertainty draws only at the level of the country.

## Footnotes

1 MMR is defined as deaths per 100,000 live births. In sub-Saharan Africa in 2015 it was 547; in the US in 1936 it was 555.

2 Duflo (2012) notes: “other than pre-birth and in early childhood, women are most likely to be missing relative to men in childbearing years.” Of the 6 million missing women each year, 21% are in their reproductive years (Wong, 2012).

3 Our estimates show that GDP growth is MMR-reducing, albeit less effective than implementation of gender quotas. Our purpose is to highlight that there remains considerable variation in MMR conditional upon income.

4 In this period, MMR increased in a few countries, including the United States (MacDorman et al., 2016), which has the highest MMR among developed countries (Kassebaum et al., 2016). In 2015, MMR was 26.4 in the USA compared with 9.2 in the UK and 4.4 in Sweden per 100,000 live births. Research investigating the potential for women in politics to reverse this trend is merited. In a recent tweet based on Mann et al. (2018), Amitabh Chandra makes a point similar to ours concerning allocative inefficiency related to interventions around women’s health, see [Twitter: Amitabh Chandra](https://twitter.com/amitabhchandra2/status/1059123396552867840) (stored for posterity [here](http://www.damianclarke.net/resources/tweetChandra_Mmort.png)).

5 In the few causal studies available, Pettersson-Lidbom (2014) estimates that a 1% increase in the share of midwife-assisted home-births decreased MMR by 2% in 19^th^ century Sweden, and Anderson et al. (2016) document that occupational licensing of midwives reduced MMR by 6-8 percent in the US in the early 20^th^ century.

6 Our analysis period is the same, 1990–2015. However, the estimated declines in this paper emerge from the 22 countries mandating quotas.

7 See, for instance, the review by Pande and Ford (2012), who discuss the cross-country implementation of quotas but provides evidence emerging only from implementation of local government quotas in India.

8 Although see Bhalotra, Venkataramani and Walther (2018) for contrasting evidence showing increases in fertility and reductions in labor force participation.

9 The countries implementing quotas are: Afghanistan, Algeria, Bangladesh, Burundi, China, Djibouti, Eritrea, Haiti, Iraq, Jordan, Kenya, Morocco, Niger, Pakistan, Rwanda, Saudi Arabia, South Sudan, Sudan, Swaziland, Tanzania, Uganda and Zimbabwe. Samoa implemented quotas in 2016 after the MMR data became available, and we do not have data for Kosovo, Somalia and Taiwan, which have implemented quotas. Uganda is the only country which reserved seats before 1990, in 1989.

10 Mean MMR in the global sample is 233 per 100,000 births, with range, 3 to 2890. The width of the range demonstrates the potential for reduction. Notice that mean adult male mortality is 238 per 1,000 male population, with range 58.8 to 689 (Table A1).

11 If anything, the incidence and death rates from TB are higher among men than women: “In 2017 close to 6 million adult men contracted TB and around 840,000 died from it. This compares with an estimated 3.2 million adult women who fell ill and almost half a million who died from TB” (WHO, September 2018).

12 The estimates are not sensitive to shrinking the lags since impacts endure.

13 The median (mean) gender quota is 21% (20%). The estimated impacts of quotas on the proportion of women in parliament are smaller than the entire size of quotas. In quota implementing countries the pre-quota share of women in parliament was not always zero, the average was 7.9%, rising to 20.9% post-quota (median: 6.2 and 21.0%). Taking all countries, the mean was 14.1%, median 11.5% (see Figures A5 and B2 for full distributions). See Figure B3 for temporal variation by country. In Rwanda we see a jump in line with quota legislation but from a high baseline, while Djibouti shows a sharp jump from zero to quota attainment. In some countries, it took time from quota passage until fulfillment. In Niger, for instance, the quota was in 2000 but the next election in 2004.

14 Democracy raises women’s share when the score is at least 6 on a scale 0–10, and directly impacts MMR when the score is 9 or 10.

15 There is no clear agreement on the precise definition of democracy. Besley and Kudamatsu (2008) discuss two particular cut-offs based on democracy scores issued in Polity IV. According to the often used definition where any non-zero Polity-IV score is classified as democratic, 7 of 22 quota adopting countries were non-democratic in the 5 years pre- and post-quota adoption (China, Eritrea, Morocco, Rwanda, Saudi Arabia, Swaziland and Uganda). If the more demanding cut-off of a polity-IV score greater than 5 is used, 14 of 22 quota adopting countries would be classified as non-democratic for this period.

16 Women tend to show more intrinsic motivation than men (Folbre, 2012). In the political domain, Baskaran et al. (2018) argue that women legislators in India exhibit more intrinsic rather than extrinsic motivation based upon comparing male and female legislator performance in swing vs non-swing constituencies in a sample of close elections between men and women.

17 We note that the instrument does not always pass a weak instrument test, but present these as ancillary estimates.

18 Since progress on women’s rights is of particular interest, we investigated this further. Using recently collated data on women’s political, social and economic rights (Cingranelli, Richards and Clay, 2013), we estimate a different event study, regressing the rights index on a series of year dummies, defined for a small window around the (country-specific) date of quota legislation. Consistent with quotas being part of the index of political rights, we observe a trend break in this index. However, we see no evidence of trend breaks in women’s economic and social rights coincident with the date of quota legislation. Thus it does not seem that the passage of gender quotas is capturing secular generic progress in women’s rights.

19 The maternal mortality data are published by the MMEIG along with 80% uncertainty intervals associated with each point estimate. In describing the modeling procedure, the authors note “We computed 80% uncertainty intervals (UIs) for the MMR and all related outcomes using the 10th and 90th percentiles of the posterior distributions. …We report 80% UIs rather than 95% UIs because of the substantial uncertainty inherent in maternal mortality outcomes: intervals based on higher uncertainty levels quickly lose their ability to present meaningful summaries of a range of likely outcomes.” (Alkema et al., 2016, p. 1250)

20 In each of the two types of distributions we allow for the possibility of either assuming full correlation in uncertainty by country or not. The corrections by country assume full correlation in uncertainty within a country over time. Where not by country, the estimator assumes no correlation within a country over time. The triangular corrections re-sample from MMR so that coverage respects the full 80% uncertainty interval suggested by the MMEIG. The normal corrections re-sample so that 80% of re-samples fall within the 80% uncertainty interval reported by the MMEIG. Both of these inference procedures are implemented assuming no correlation by country, and then assuming full correlation in uncertainty by country.

21 The political identity literature tends to refer to leader preferences and there is some evidence that leaders influence not only citizens of their identity but the wider population in accordance with their preferences (see Bhalotra, Clots-Figueras and Iyer (2018); Bassi and Rasul (2017)). However, in democratic regimes, an alternative possibility is that women leaders implement women-friendly policies for strategic reasons, namely, to incentivize women voters to re-elect them.

22 In addition to having stronger preferences over health, women may be more effective at delivering health if, because of their greater involvement in child-bearing and caring for the sick, health is more salient in their minds or they have more information on how it can be addressed.

23 Infant mortality is a widely used proxy for public health in the earlier stages of development. These results may be further influenced by the tendency for women to dedicate more resources toward children than men, possible evolutionary roots of which lie in paternity uncertainty (Cox, 2007).

24 The evidence is suggestive rather than conclusive. First, mortality rates by cause of death and age for many developing countries are inadequate, for instance, see Klasen and Vollmer (2013). The MMR data we use were generated by a major multi-UN task force, no analogue of which has been appointed to create estimates for other categories of mortality. If there were more measurement error in measuring TB or male mortality than in measuring MMR and if that measurement error were classical, then the coefficients for TB and male mortality would be noisier. This said, as we also find that they are smaller, or less negative, this alone is unlikely to explain away our finding of differentially large effects on MMR. Second, one needs to be careful comparing changes delivered by gender quotas for conditions with different treatability. For instance, the injuries and accidents that contribute to adult male mortality may be harder for policy to address than the reproductive health services that contribute to maternal mortality.

25 For completeness, we also present estimates for adult female mortality. Visual inspection of the plot suggests a sharper response of female than of male adult mortality, but it is not significant. This does not surprise us because any policies targeting causes of adult mortality among women other than MMR (TB, accidents, etc.) are not easily targeted to women. The share of MMR in female adult mortality varies in the sample, from 0.002 (Finland, Greece and Poland) to 0.343 (Niger).

26 We do not measure outcomes for every domain of health spending, so in principle some unmeasured outcome may have suffered. However, as discussed in the section 4, adult male mortality is a natural analogue to MMR, infant mortality is a marker of population health, and TB remains an important cause of illness and death for men and women in poor countries.

27 Miller (2008) cites evidence from historical studies that women conducted door to door information campaigns in early 20^th^ century America, to encourage families to boil drinking water, and this contributed to sharp reductions in infant mortality. Baskaran et al. (2018) show that women legislators in India are more likely to complete an allocated village road building project in their constituency than are male legislators. They also show that women are less corrupt than men and that, in contrast to men, they act to raise economic activity in their constituencies even when they have won in a non-swing constituency. They argue that, if economic growth is the common currency in which costs are evaluated, then having women instead of men in government imposes no economic cost. Beaman et al. (2009) show that quotas for women in village council leadership in India led to raised aspirations and higher educational investments in girls. Bhalotra and Clots-Figueras (2014) show that breastfeeding rates in Indian districts increase when women legislators are elected from the district.

